# Effect of red blood cell transfusion strategies on ICU-acquired infection in patients with sepsis: A target trial emulation using the Medical Information Mart for Intensive Care IV database

**DOI:** 10.64898/2026.08.14.26360449

**Authors:** Shodai Yoshihiro, Yuki Kataoka, Mitsuaki Nishikimi, Nobuaki Shime, Hiroaki Matsuo

## Abstract

**Purpose:** To estimate the per-protocol effect of red blood cell (RBC) transfusion strategies on ICU-acquired infection in critically ill adults with sepsis using a target trial emulation framework. We evaluated whether restrictive strategy and liberal strategy, defined by hemoglobin (Hgb) thresholds, differ in their effect on ICU-acquired infection during ICU stay.

**Methods:** We conducted a target trial emulation using the MIMIC-IV database and included adults who met Sepsis criteria at ICU admission. Clones were assigned to restrictive or liberal transfusion strategies. Under the restrictive strategy, RBC transfusion was permitted only when Hgb was ≤7.0 g/dL, whereas under the liberal strategy, transfusion was permitted when Hgb was >7.0 g/dL. The primary outcome was the first ICU-acquired infection occurring at least 72 hours after ICU admission. Per-protocol effects were estimated using a clone-censor-weight approach with a marginal structural model. A parametric g-formula was used as a complementary analysis that jointly modeled ICU discharge and ICU mortality as competing events to derive strategy-specific 28-day cumulative incidences and risk differences.

**Results:** Among 4,013 eligible ICU stays, the liberal-versus-restrictive comparison provided little evidence of a difference in the risk of ICU-acquired infection (adjusted conditional OR, 0.954; 95% CI, 0.797–1.142). In the complementary g-formula analysis, the 28-day risk difference for the liberal versus restrictive comparison was −0.02 percentage points (95% CI, −0.15 to 0.11), consistent with the primary analysis. Findings were generally robust across prespecified subgroup and sensitivity analyses.

**Conclusion:** In this target trial emulation of adults with sepsis, we observed no clinically meaningful difference in ICU-acquired infection between RBC transfusion strategies defined by hemoglobin thresholds.

## 1. Introduction

Sepsis is a life-threatening syndrome characterized by organ dysfunction caused by a dysregulated host response to infection [1]. In addition to source control and antimicrobial therapy, resuscitative interventions are used to restore the balance between tissue oxygen delivery and demand. Red blood cell (RBC) transfusion is one such intervention intended to augment oxygen-carrying capacity during resuscitation.

However, the optimal RBC transfusion strategy for adults with sepsis has not been established. International clinical practice guidelines recommend a restrictive strategy based on hemoglobin (Hgb) thresholds in patients with sepsis [2, 3]. This recommendation is supported in part by randomized controlled trials (RCTs) showing no increase in mortality with the restrictive strategy compared with the liberal strategy [4, 5], as well as by considerations of resource utilization, cost-effectiveness, and equity in health care delivery. Nevertheless, the available evidence remains insufficient to establish noninferiority with adequate precision, as the required information size has not been reached [6]. Moreover, any potential benefit of RBC transfusion must be weighed against the risk of adverse events, including acute lung injury, circulatory overload, and immunomodulatory effects.

In particular, the relation between RBC transfusion and infection risk remains incompletely understood. Pediatric RCTs have suggested that the restrictive strategy may reduce infection risk [7, 8], whereas evidence from adult RCTs remains inconclusive [4, 5]. An observational study also suggested that RBC transfusion may be associated with an increased risk of infection [9]. However, that study did not adequately address time-dependent confounding or immortal time bias, limiting causal interpretation.

Therefore, we used target trial emulation with a marginal structural model (MSM) to estimate the per-protocol effect of RBC transfusion strategies on intensive care unit (ICU)-acquired infection in patients with sepsis.

## 2. Methods

### 2.1 Study Design and Data

We emulated a hypothetical pragmatic randomized trial (a “target trial”) in which septic patients in the ICU were assigned to three groups: a restrictive strategy in which transfusions were permitted at Hgb levels ≤7.0 g/dL, the liberal strategy in which transfusions were permitted at Hgb levels >7.0 g/dL, and a no-transfusion strategy. The target trial was based on a previous RCT [4], which compared lower versus higher Hgb thresholds for RBC transfusion in patients with septic shock and found no significant difference in 90-day mortality between the groups. The detailed specification of the target trial and its corresponding observational emulation are provided in **Supplementary Table 1**.

We used the Medical Information Mart for Intensive Care IV (MIMIC-IV) database, version 3.1. MIMIC-IV is a large, publicly available, single-center database comprising de-identified comprehensive electronic health records of patients admitted to the ICUs at Beth Israel Deaconess Medical Center in Boston, Massachusetts, United States, between 2008 and 2023 [10]. We had access to the de-identified source tables within MIMIC-IV required to define the study cohort, exposures, outcomes, and covariates, and cohort selection as well as construction of the analytic dataset were performed by the investigators. To report this study, we followed the RECORD statement [11], the TARGET statement [12], and the reporting guideline for multiple imputation by chained equations (MICE) [13]. We provide the RECORD checklist in the **Supplementary Table**.

In the primary analysis, an MSM was used to estimate the per-protocol causal contrast as a cause-specific odds ratio (OR) for ICU-acquired infection during ICU survival. In the secondary analysis, a parametric g-formula was used to model day-by-day transitions among infection, ICU discharge, and death and to simulate counterfactual trajectories under each strategy, yielding strategy-specific 28-day cumulative incidence functions (CIFs) and corresponding risk differences. The MSM provided a concise relative effect estimate under ideal adherence, whereas the g-formula explicitly incorporated competing events and time-varying covariates to estimate intervention-specific absolute risks over time.

### 2.2 Eligibility Criteria

We identified adults (aged ≥18 years) admitted to the ICU who met the Sepsis criteria at ICU admission. Sepsis was defined by the simultaneous presence of suspected infection and accompanying organ dysfunction, using a clinical surveillance definition adapted from Rhee et al. [14] and the official MIMIC-IV implementation.

Suspected infection was identified by the co-occurrence of microbiological culture sampling and initiation of intravenous antibiotic therapy within clinically relevant time windows around ICU admission. Accompanying organ dysfunction was defined as the presence of at least one of six clinical criteria (the cardiovascular, respiratory, renal, hepatic, coagulation, or metabolic systems) [14]. The specific thresholds and reference value definitions for each criterion are detailed in **Supplementary Table 2**.

The following patients were excluded: (1) those who died or were discharged from the ICU within 48 hours of admission; (2) patients with massive hemorrhage [15, 16]; (3) patients with hematologic diseases; and (4) patients receiving comfort measures only [14, 17]. If the interval between ICU stays was less than 48 hours, the subsequent ICU admission was excluded from the analysis. The complete lists of diagnosis codes are provided in **Supplementary Table 2**.

The study size was determined by the number of eligible ICU stays available in MIMIC-IV v3.1 after application of the prespecified inclusion and exclusion criteria.

### 2.3 Treatment Strategies

#### Definition of Strategies

Patients were classified according to one of three mutually exclusive RBC transfusion strategies based on their daily Hgb levels and transfusion status: (1) the restrictive strategy, in which RBC transfusion was permitted only when Hgb fell to ≤7.0 g/dL; (2) the liberal strategy, in which RBC transfusion was permitted at higher Hgb thresholds (Hgb >7.0 g/dL); and (3) a no-transfusion strategy. Protocol adherence was assessed daily. In brief, for the restrictive strategy, transfusion when Hgb was >7.0 g/dL was considered a protocol deviation; for the no-transfusion strategy, any RBC transfusion was considered a deviation. The full operational definitions for all three strategies are provided in **Supplementary Table 2**.

We followed up patients for a maximum of 28 days from time zero. Follow-up for each patient (or clone) was terminated at the earliest occurrence of: deviation from the assigned transfusion strategy, ICU-acquired infection, death, ICU discharge, or completion of the 28-day observation period.

### 2.4 Definition of Outcomes

#### Primary Outcome

The primary outcome was the first episode of ICU-acquired infection, defined as a new infection occurring at least 72 hours after ICU admission for sepsis [18].

ICU-acquired infection was identified using an established clinical surveillance definition [14]. Two restrictions were applied to isolate genuinely ICU-acquired, de novo infections from the index sepsis episode and from ongoing antimicrobial management. First, infections with onset more than 72 hours after ICU admission were eligible as counted. Second, an antibiotic start required no administration of a different intravenous antibiotic in the preceding 24 hours. The specific Item IDs and detailed operational definitions are provided in **Supplementary Table 3**.

#### Competing Events

Death and ICU discharge were treated as competing events that precluded the occurrence of ICU-acquired infection. The operational definitions of these events are provided in **Supplementary Table 3**.

#### Observation Period

The maximum observation period was 28 days from time zero (ICU admission). Events occurring beyond Day 28 were administratively censored. Within this window, follow-up for each patient (or clone) ended at the earliest of the primary outcome, death, ICU discharge, strategy deviation (artificial censoring), or the 28-day boundary.

### 2.5 Covariates

We drew two directed acyclic graphs (DAGs) to identify potential confounders of the relationship between transfusion strategy and ICU-acquired infection (**Supplementary Figure 1**).

The following time-fixed baseline covariates were measured before the first transfusion: age at ICU admission, baseline Sequential Organ Failure Assessment (SOFA) score, baseline Hgb level, serum albumin, body mass index (BMI), and five comorbidity indicators (congestive heart failure, chronic pulmonary disease, renal disease, mild or severe liver disease, and rheumatic disease). Two time-varying covariates were updated daily: daily Hgb and daily SOFA score. Data sources and operational definitions for all covariates are provided in **Supplementary Table 4**.

### 2.6 Statistical Analysis

We evaluated the per-protocol effect of three transfusion strategies on ICU-acquired infection. In the primary analysis, we used a clone–censor–weight approach embedded within a MSM to estimate the effect as a weighted, covariate-adjusted conditional OR with 95% confidence intervals (CIs) [19–21].

Clones were created using baseline covariates, and follow-up was discretized into daily intervals. Within the MSM framework, we modeled adherence to the assigned strategy over time by fitting a logistic regression model at each daily time point to estimate the probability of remaining adherent, conditional on baseline covariates and time-varying confounders. We then calculated the inverse probability of censoring weighting (IPCW) to account for informative censoring due to deviations from the assigned strategy. Since individuals were duplicated into strategy-specific clones and were therefore identical at baseline across strategies, inverse probability of treatment weighting was not applied. A detailed description of the MSM implementation, including model specifications and weighting procedures, is provided in **Supplementary Methods**.

#### Additional Effect Measures and Sensitivity Analyses

Although the conditional OR from the primary MSM estimates the effect for individuals with a given set of baseline covariates, we additionally derived population-averaged effect measures that may be more directly interpretable in clinical practice. Using marginal standardization, we estimated the predicted probability of ICU-acquired infection under each strategy for each patient and averaged these probabilities over the observed baseline covariate distribution. The marginal approach summarizes the average causal effect across the study population and allows estimation of both marginal ORs and absolute risk differences with 95% CIs.

We conducted several pre-specified sensitivity analyses to evaluate the robustness of our findings. First, we evaluated a model that removed baseline covariates from both the IPCW numerator and the MSM outcome model (No baseline vars). Second, we modified the timing of artificial censoring for protocol deviations to apply on the same day of the deviation rather than the next day (Deviation Timing). Third, we performed a landmark analysis defining Day 1 as time zero, including pre-Day 1 RBC transfusion as an additional baseline covariate to address potential immortal time bias (Landmark Day 1). Fourth, we examined sensitivity to the IPCW truncation thresholds by applying the 95th and 99th percentiles (Truncation 95% and 99%). Fifth, we performed a complete case analysis restricted to patients with no missing required baseline or time-varying covariates (Complete Case). Finally, we ascertained hospital-acquired infection through Day 28 regardless of ICU discharge to extend the observation window (28-Day HAI Observation).

#### Subgroup Analysis

As a pre-specified supplementary analysis, we repeated the clone–censor–weight and MSM procedure after restricting the population to patients with baseline SOFA score >5 [22].

#### Secondary Analysis: Parametric g-formula

As a complementary analysis, we used the parametric g-formula to estimate the 28-day cumulative incidence of ICU-acquired infection under each transfusion strategy [23]. In the MSM, competing events were handled using inverse probability weighting, whereas the g-formula explicitly models them jointly with time-varying covariates to directly estimate cumulative incidence under each intervention. We fit parametric models for time-varying covariates, transfusion status, and competing outcomes (ICU-acquired infection, ICU discharge, and in-ICU death) and implemented a Monte Carlo simulation to generate counterfactual event trajectories through day 28. We derived CIFs and between-strategy risk differences from the simulated data, with uncertainty quantified using bootstrap resampling and multiple imputation. Full model specifications and simulation procedures are provided in the **Supplementary Methods**.

#### Missing Data

Missing values in baseline and time-varying covariates were handled using MICE [24]. The imputation model included all analysis variables, the outcome, and auxiliary variables expected to be associated with missingness. Results were pooled across imputed datasets using Rubin’s rules. Across 20 imputed datasets, the distributions of imputed values were visually confirmed with the observed distributions via kernel density plots (**Supplementary Figure 2**).

The imputation procedures and number of datasets are described in the **Supplementary Methods**.

#### Software

All analyses were conducted using R (version 4.4.1; R Foundation for Statistical Computing). The clone-censor-weight procedure and inverse probability weighting were implemented following the code provided in Chapter 12 of Hernán and Robins [25]. The parametric g-formula was implemented as custom code whose algorithm design, particularly the handling of history functions and competing events, was kept consistent with the R package gfoRmula [26], while being optimized for the data structure of the present study. Additional key packages included splines, sandwich, lmtest, and mice. Statistical significance was assessed at a two-sided α level of 0.05. Claude Opus and Gemini 3 were used to assist with code writing, debugging, and refinement for data-processing and modeling scripts. The codes and algorithms used for cohort selection, variable construction, and statistical analysis are available in a GitHub repository [27]. The raw data are available from the MIMIC-IV database, subject to its access requirements.

## 3. Results

### 3.1 Study Population and Baseline Characteristics

Of 546,028 adult ICU admissions in the MIMIC-IV database, 49,484 met criteria for suspected infection and 14,925 fulfilled sepsis criteria. After applying prespecified exclusions, including 2 stays with structural data errors, the primary analytic cohort included 4,013 ICU stays (**Figure 1**), which included 101 non-first ICU stays within a single hospital admission. Among these, 60 stays (1.5%) had Hgb measurements available only during the first 24 hours after ICU admission or within the final 24-hour interval before ICU discharge and were retained in the primary analysis. Baseline characteristics at time zero are summarized in **Supplementary Table 5**. Missingness among baseline variables was highest for serum albumin (68.9%) and baseline bilirubin (55.4%). The complete-case sample comprised 111 stays. Detailed missingness patterns and imputation diagnostics are provided in **Supplementary Table 5** and **Supplementary Figure 2**.

**Figure 1:**
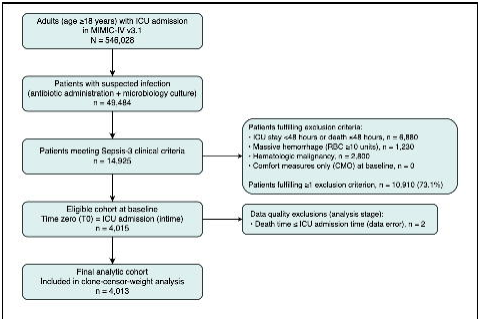
Flow diagram of participant selection for the Sepsis analytic cohort. The flow diagram shows the sequential application of inclusion and exclusion criteria to the source population of 546,028 adult ICU stays in MIMIC-IV v3.1, leading to the eligible cohort at baseline (n = 4,015 ICU stays) and the final analytic cohort (n = 4,013 ICU stays) after data quality exclusions.

### 3.2 Outcome Events

Under the primary definition, 152 ICU-acquired infection events were identified, with a median time from ICU admission to event of 7.14 days (IQR, 4.43–10.51). Under the any-event definition, 426 first valid infection events were identified, with a median time from ICU admission to event of 4.51 days (IQR, 3.54–6.64).

Under the de novo HAI over the 28-day follow-up, 222 events were identified, with a median time from ICU admission to event of 7.92 days (IQR, 5.00–11.69). Under the any-event definition, 576 first valid HAI events were identified, with a median time from ICU admission to event of 4.90 days (IQR, 3.67–7.83).

### 3.3 Follow-up

At time zero, each of the 4,013 eligible ICU stays was cloned across the three transfusion strategies, yielding 4,013 clones per strategy (restrictive strategy, liberal strategy, and no-transfusion strategy). During follow-up, artificial censoring for protocol deviation occurred in 654 of 4,013 restrictive strategy clones (16.3%), 1,168 of 4,013 liberal strategy clones (29.0%), and 673 of 4,013 no-transfusion strategy clones (16.8%). Total follow-up time was 19,457 person-days under the restrictive strategy, 16,161 person-days under the liberal strategy, and 19,303 person-days under no-transfusion strategy.

### 3.4 Transfusion Patterns

The proportion of clones that received any RBC transfusion was 1.1% under the restrictive strategy and 8.6% under the liberal strategy. Among recipients of RBC transfusion, temporal transfusion patterns are summarized in **Supplementary Table 6, Supplementary Figure 3 and Supplementary Figure 4**.

### 3.5 Main Outcomes

In the primary MSM, the adjusted conditional OR for ICU-acquired infection during ICU stay was 0.954 (95% CI, 0.797–1.142; p = 0.609) for the liberal-versus-restrictive comparison (**Figure 2**). For the no-transfusion-versus-restrictive comparison, the corresponding adjusted conditional OR from the primary MSM was 0.994 (95% CI, 0.926–1.067; p = 0.864), indicating little difference in the estimated per-protocol effect. (**Figure 2**).

**Figure 2:**
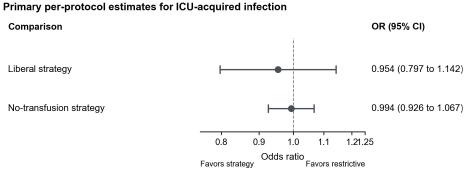
Primary per-protocol estimates for ICU-acquired infection during the ICU stay. Odds ratios and 95% confidence intervals were estimated using the primary marginal structural model. The restrictive transfusion strategy was the reference. Odds ratios below 1 favor the strategy shown.

After marginal standardization to the observed baseline covariate distribution, the estimated population-averaged effect for the liberal-versus-restrictive comparison was a marginal OR of 0.955 (95% CI, 0.799–1.141; p = 0.608) and a marginal risk difference of −0.02 percentage points (95% CI, −0.12 to 0.07; p = 0.630). These marginally standardized estimates were similar in direction and magnitude to the conditional MSM estimates. Stabilized IPCW weights in the primary analysis had a mean of approximately 0.999 (SD, 0.086) and a maximum of 1.259 after truncation at the 97.5th percentile (effective sample size ratio > 99%), suggesting no evidence of positivity violations or extreme weight instability.

In a subgroup analysis restricted to patients with higher disease severity (baseline SOFA score >5; n = 2,568; 63.9%), the estimated marginal effect for the liberal-versus-restrictive comparison was a marginal OR of 0.941 (95% CI, 0.770–1.152; p = 0.557) and a risk difference of −0.04 percentage points (95% CI, −0.18 to 0.10; p = 0.582). Stabilized IPCW weights in this subgroup had a mean of approximately 0.997 (SD 0.092) and a maximum of 1.266 (effective sample size ratio >99%); estimates were similar across weight truncation thresholds (**Supplementary Table 7**).

Across prespecified sensitivity analyses, the estimated effect for the liberal-versus-restrictive comparison remained directionally similar to that of the primary analysis. These analyses included models that removed baseline covariates from the IPCW numerator and the MSM outcome model (OR, 1.023; 95% CI, 0.787–1.330; p = 0.865), altered the timing of artificial censoring, used a Day-1 landmark, applied alternative weight-truncation thresholds, or were restricted to complete cases (**Supplementary Table 8**). A similar pattern was observed when the ascertainment of ICU-acquired infection was extended through Day 28 irrespective of ICU discharge, with estimates close to the null across all three strategy comparisons (Supplementary Table 8).

### 3.6 Secondary Analyses

In the secondary parametric g-formula analysis pooled across 20 imputed datasets, the estimated 28-day cumulative incidence of ICU-acquired infection under each strategy was 2.16% (95% CI, 2.07–2.25) for the restrictive strategy, 2.14% (95% CI, 2.06–2.23) for the liberal strategy, and 2.22% (95% CI, 2.12–2.32) for the no-transfusion strategy. The corresponding 28-day risk differences were −0.02 percentage points (95% CI, −0.15 to 0.11) for the liberal-versus-restrictive comparison and +0.06 percentage points (95% CI, −0.08 to 0.20) for the no-transfusion-versus-restrictive comparison (**Figure 3; Supplementary Table 9 and 10**; **Supplementary Figure 5 and 6**).

**Figure 3:**
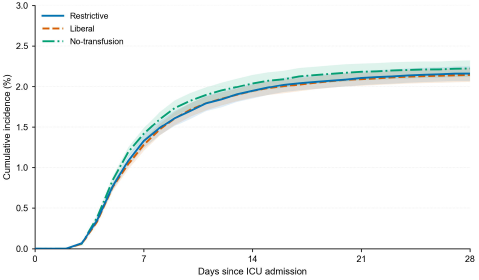
Estimated cumulative incidence of ICU-acquired infection under each transfusion strategy. Curves show cumulative incidence estimates obtained from the secondary parametric g-formula analysis pooled across 20 imputed datasets. Shaded areas indicate pointwise 95% confidence intervals. Estimates are shown through Day 28 after ICU admission.

## 4. Discussion

In this target trial emulation of three RBC transfusion strategies among critically ill adults with sepsis, we estimated the per-protocol effect of each strategy on ICU-acquired infection using 4,013 ICU stays from the MIMIC-IV database. With the restrictive strategy as the reference, the estimated effect of the liberal strategy provided little evidence of a difference in the risk of ICU-acquired infection (adjusted conditional OR, 0.954; 95% CI, 0.797–1.142). Similarly, the estimated effect for the no-transfusion strategy versus restrictive strategy comparison was close to the null, with little evidence of a meaningful difference in ICU-acquired infection risk (adjusted conditional OR, 0.994; 95% CI, 0.926–1.067). These findings remained consistent in the prespecified subgroup with higher baseline illness severity (SOFA > 5), and and across prespecified sensitivity analyses. In a complementary analysis, we used the g-formula to estimate the strategy-specific 28-day cumulative risk of infection while explicitly accounting for ICU discharge and in-ICU death as competing events. This analysis also showed no difference in absolute risk for the liberal-versus-restrictive comparison (risk difference, −0.02%; 95% CI, −0.15 to 0.11), supporting the consistency of inference across the two causal approaches.

### 4.1 Interpretation of Findings

To our knowledge, no previous study has directly compared ICU-acquired infection across prespecified RBC transfusion strategies in critically ill adults with sepsis. Transfusion-related adverse events have not been adequately evaluated in primary studies of adults [4, 5], although studies in pediatric patients with sepsis suggested that the restrictive strategy may reduce infection-related complications [7, 8]. Consequently, guideline recommendations for RBC transfusion in critically ill adults with sepsis have limited direct evidence regarding infection-related harms [2, 3]. In particular, prior observational studies reported an association between observed exposure to RBC transfusion and an increased risk of infection [9]. Although Dupuis et al. also addressed time-varying confounding, their analysis estimated the effect of observed transfusion exposure rather than the effect of prespecified transfusion strategies [9]. In contrast, our target trial emulation estimated the per-protocol effect of explicit strategies by aligning eligibility, strategy assignment, and follow-up at a common time zero using the clone–censor– weight approach. This design reduces biases that can arise when these elements are not synchronized. By addressing several analytic limitations of prior studies, our study suggests that the difference in ICU-acquired infection risk between transfusion strategies may be smaller than previously implied.

In our study, infection risk was not higher under the liberal strategy than under the restrictive strategy. This occurred even though the liberal strategy entailed greater transfusion exposure. At first glance, this appears to conflict with prior work. Pediatric RCTs suggested that the restrictive strategy may reduce infection [7, 8]. An observational cohort reported that greater observed transfusion exposure was associated with increased infection risk [9]. One explanation for this apparent discrepancy is dose dependence. That is, the effect of transfusion on infection may be dose dependent rather than uniform. Experimental and translational work indicates that transfusion can elicit both immunosuppressive and immune-activating responses [28]. The net effect at a given level of exposure may therefore reflect the balance between these opposing mechanisms. It may not reflect a uniformly immunosuppressive effect. Consistent with this, a dose-response meta-analysis suggested that the relation between transfusion exposure and infection risk is nonlinear [29]. If the relationship is dose dependent, the size of the between-strategy difference in transfusion exposure becomes critical. In the pediatric RCT, this difference was substantial (median transfused volume 7.6 vs 15.7 mL/kg, approximately 8 mL/kg) [8]. In our cohort, it was less than 1 unit (9.3 vs 11.0 mL/kg). At these comparatively small doses, the net immunomodulatory effect may have been more favorable with respect to infection under the liberal strategy. This interpretation nonetheless remains speculative. The estimated reduction in infection risk under the liberal strategy was also small. Concern about ICU-acquired infection alone may therefore be insufficient to justify the restrictive strategy in this setting.

### 4.2 Strengths and Limitations

A major strength of this study is the explicit target trial emulation framework. The clone–censor– weight approach aligned eligibility, strategy assignment, and time zero, and IPCW was used to account for censoring induced by deviations from the assigned strategy. We also compared results across complementary causal methods, including an MSM and the parametric g-formula, which improved the transparency of the assumptions and the stability of the substantive interpretation.

Some limitations should also be considered. First, this study was based on 15 years of accumulated data from a single center, and secular trends, including changes in infection control policies and clinical practice over time, may not have been fully adjusted for. Therefore, the external validity of the findings should be interpreted with caution.

Second, despite adjustment for measured baseline and time-varying confounders, residual confounding remains possible, particularly from factors that were unavailable or incompletely captured in the database. Although we used the DAG to select measured confounders, we could not account for potentially important features of the intervention and care process such as storage age or other product characteristics.

## 5. Conclusions

In our target trial emulation, no clinically meaningful difference in ICU-acquired infection during the ICU stay was observed between RBC transfusion strategies defined by Hgb thresholds. Accordingly, among critically ill adults with sepsis, concern about ICU-acquired infection alone may play only a limited role when clinicians weigh the overall effects of RBC transfusion strategies. Further evaluation using multicenter data is warranted in settings where transfusion exposure differs more substantially between strategies.

## Data Availability

Data availability: This study used de-identified data from the MIMIC-IV database, version 3.1, which is available through PhysioNet to credentialed users who complete the required training and sign the Data Use Agreement. The authors are not permitted to redistribute the raw data. The code and algorithms used for cohort selection, variable construction, and statistical analysis are available in the GitHub repository cited in the manuscript.

https://github.com/Yoshihiro-glitch/Target-Trial-Emulation_Transfusion-Strategy-public

## 6. Acknowledgments

## Conflict of Interest

The authors have no conflicts of interest to declare.

The authors used M365 Copilot during manuscript revision to assist with English-language editing and improvement of clarity, and used Claude Opus and Gemini 3 to assist in drafting code. The authors reviewed and verified all outputs and take full responsibility for the content of this manuscript.

## Abbreviations

BMI: body mass index
CIs: confidence intervals
CIF: cumulative incidence function
DAG: directed acyclic graph
Hgb: hemoglobin
ICU: intensive care unit
IPCW: inverse probability of censoring weighting
MICE: multiple imputation by chained equations
MIMIC-IV: Medical Information Mart for Intensive Care IV
MSM: marginal structural model
OR: odds ratio
RBC: red blood cell
RCT: randomized controlled trial
SOFA: Sequential Organ Failure Assessment

