## Supplementary Appendix for "Effect of red blood cell transfusion strategies on ICU-acquired infection in patients with sepsis: A target trial emulation using the Medical Information Mart for Intensive Care IV database"

### The RECORD statement – checklist of items, extended from the STROBE statement, that should be reported in observational studies using routinely collected health data.

|  | **Item No.** | **STROBE items** | **Location　in manuscript where items are reported** | **RECORD items** | **Location in manuscript where items are reported** |
| --- | --- | --- | --- | --- | --- |
| **Title and abstract** | | | | | |
|  | 1 | (a) Indicate the study’s design with a commonly used term in the title or the abstract (b) Provide in the abstract an informative and balanced summary of what was done and what was found | 1, 2 | RECORD 1.1: The type of data used should be specified in the title or abstract. When possible, the name of the databases used should be included.  RECORD 1.2: If applicable, the geographic region and timeframe within which the study took place should be reported in the title or abstract.  RECORD 1.3: If linkage between databases was conducted for the study, this should be clearly stated in the title or abstract. | 1, 2 |
| **Introduction** | | | | | |
| Background rationale | 2 | Explain the scientific background and rationale for the investigation being reported | 3 |  |  |
| Objectives | 3 | State specific objectives, including any prespecified hypotheses | 3 |  |  |
| **Methods** | | | | | |
| Study Design | 4 | Present key elements of study design early in the paper | 3 |  |  |
| Setting | 5 | Describe the setting, locations, and relevant dates, including periods of recruitment, exposure, follow-up, and data collection | 4 |  |  |
| Participants | 6 | *(a) Cohort study* - Give the eligibility criteria, and the sources and methods of selection of participants. Describe methods of follow-up  *Case-control study* - Give the eligibility criteria, and the sources and methods of case ascertainment and control selection. Give the rationale for the choice of cases and controls  *Cross-sectional study* - Give the eligibility criteria, and the sources and methods of selection of participants  *(b) Cohort study* - For matched studies, give matching criteria and number of exposed and unexposed  *Case-control study* - For matched studies, give matching criteria and the number of controls per case | 5-8 | RECORD 6.1: The methods of study population selection (such as codes or algorithms used to identify subjects) should be listed in detail. If this is not possible, an explanation should be provided.  RECORD 6.2: Any validation studies of the codes or algorithms used to select the population should be referenced. If validation was conducted for this study and not published elsewhere, detailed methods and results should be provided.  RECORD 6.3: If the study involved linkage of databases, consider use of a flow diagram or other graphical display to demonstrate the data linkage process, including the number of individuals with linked data at each stage. | 5-8 |
| Variables | 7 | Clearly define all outcomes, exposures, predictors, potential confounders, and effect modifiers. Give diagnostic criteria, if applicable. | e-method | RECORD 7.1: A complete list of codes and algorithms used to classify exposures, outcomes, confounders, and effect modifiers should be provided. If these cannot be reported, an explanation should be provided. | e-method  Reference [ ] |
| Data sources/ measurement | 8 | For each variable of interest, give sources of data and details of methods of assessment (measurement).  Describe comparability of assessment methods if there is more than one group | 7, 9 |  |  |
| Bias | 9 | Describe any efforts to address potential sources of bias |  |  |  |
| Study size | 10 | Explain how the study size was arrived at | 6 |  |  |
| Quantitative variables | 11 | Explain how quantitative variables were handled in the analyses. If applicable, describe which groupings were chosen, and why | 8-10 |  |  |
| Statistical methods | 12 | (a) Describe all statistical methods, including those used to control for confounding  (b) Describe any methods used to examine subgroups and interactions  (c) Explain how missing data were addressed  (d) *Cohort study* - If applicable, explain how loss to follow-up was addressed  *Case-control study* - If applicable, explain how matching of cases and controls was addressed  *Cross-sectional study* - If applicable, describe analytical methods taking account of sampling strategy  (e) Describe any sensitivity analyses | 9-12 |  |  |
| Data access and cleaning methods |  | .. |  | RECORD 12.1: Authors should describe the extent to which the investigators had access to the database population used to create the study population.  RECORD 12.2: Authors should provide information on the data cleaning methods used in the study. | 5  e-method |
| Linkage |  | .. |  | RECORD 12.3: State whether the study included person-level, institutional-level, or other data linkage across two or more databases. The methods of linkage and methods of linkage quality evaluation should be provided. | NA |
| **Results** | | | | | |
| Participants | 13 | (a) Report the numbers of individuals at each stage of the study (*e.g.*, numbers potentially eligible, examined for eligibility, confirmed eligible, included in the study, completing follow-up, and analysed)  (b) Give reasons for non-participation at each stage.  (c) Consider use of a flow diagram | 11, 12 | RECORD 13.1: Describe in detail the selection of the persons included in the study (*i.e.,* study population selection) including filtering based on data quality, data availability and linkage. The selection of included persons can be described in the text and/or by means of the study flow diagram. | 11, 12 |
| Descriptive data | 14 | (a) Give characteristics of study participants (*e.g.*, demographic, clinical, social) and information on exposures and potential confounders  (b) Indicate the number of participants with missing data for each variable of interest  (c) *Cohort study* - summarise follow-up time (*e.g.*, average and total amount) | 11, 12 |  |  |
| Outcome data | 15 | *Cohort study* - Report numbers of outcome events or summary measures over time  *Case-control study* - Report numbers in each exposure category, or summary measures of exposure  *Cross-sectional study* - Report numbers of outcome events or summary measures | 12 |  |  |
| Main results | 16 | (a) Give unadjusted estimates and, if applicable, confounder-adjusted estimates and their precision (e.g., 95% confidence interval). Make clear which confounders were adjusted for and why they were included  (b) Report category boundaries when continuous variables were categorized  (c) If relevant, consider translating estimates of relative risk into absolute risk for a meaningful time period | NA |  |  |
| Other analyses | 17 | Report other analyses done—e.g., analyses of subgroups and interactions, and sensitivity analyses | 13, 14 |  |  |
| **Discussion** | | | | | |
| Key results | 18 | Summarise key results with reference to study objectives | 14, 15 |  |  |
| Limitations | 19 | Discuss limitations of the study, taking into account sources of potential bias or imprecision. Discuss both direction and magnitude of any potential bias | 17, 18 | RECORD 19.1: Discuss the implications of using data that were not created or collected to answer the specific research question(s). Include discussion of misclassification bias, unmeasured confounding, missing data, and changing eligibility over time, as they pertain to the study being reported. | 17, 18 |
| Interpretation | 20 | Give a cautious overall interpretation of results considering objectives, limitations, multiplicity of analyses, results from similar studies, and other relevant evidence | 15 |  |  |
| Generalisability | 21 | Discuss the generalisability (external validity) of the study results | 16, 17 |  |  |
| **Other Information** | | | | | |
| Funding | 22 | Give the source of funding and the role of the funders for the present study and, if applicable, for the original study on which the present article is based | 18 |  |  |
| Accessibility of protocol, raw data, and programming code |  | .. |  | RECORD 22.1: Authors should provide information on how to access any supplemental information such as the study protocol, raw data, or programming code. | 11 |

*Reference: Benchimol EI, Smeeth L, Guttmann A, Harron K, Moher D, Petersen I, Sørensen HT, von Elm E, Langan SM, the RECORD Working Committee. The REporting of studies Conducted using Observational Routinely-collected health Data (RECORD) Statement. PLoS Medicine 2015; in press.

*Checklist is protected under Creative Commons Attribution (CC BY) license.

### Supplementary Table 1. Target Trial Specification and Its Emulation Using the MIMIC-IV Database

| Protocol Component | Target Trial | Target Trial Emulation |
| --- | --- | --- |
| **Eligibility** | Adults (≥18 years) newly admitted to the ICU who meet the Sepsis-3 criteria. | Adults aged ≥18 years who were newly admitted to the ICU and met sepsis criteria based on a clinical surveillance definition adapted from Rhee et al. and the official MIMIC-IV implementation. |
| **Treatment Strategies** | - **Restrictive strategy:** Initiate red blood cell (RBC) transfusion only if hemoglobin (Hgb) level drops to ≤7.0 g/dL.  - **Liberal strategy:** Initiate RBC transfusion at higher thresholds (e.g., Hgb 7.0–9.0 g/dL). | - **Restrictive strategy**: Permit RBC transfusion only when Hgb is ≤7.0 g/dL.  - **Liberal strategy**: Permit RBC transfusion when Hgb is >7.0 g/dL.  - **No-transfusion strategy**: Do not permit RBC transfusion during follow-up. |
| **Assignment** | Random assignment to a transfusion strategy at baseline (time zero). | At ICU admission, each eligible patient was cloned into three copies, with each clone assigned to one of the three strategies. The clone-censor-weight approach was then used to emulate random assignment under the assumption of no unmeasured confounding. |
| **Outcomes** | The first episode of ICU-acquired infection, defined as a new infection occurring more than 48 hours after ICU admission. | The outcome definition was identical to that specified for the target trial. |
| **Follow-up** | Starts at baseline (time zero, defined as ICU admission) and ends at the first occurrence of: death, ICU-acquired infection, deviation from the assigned strategy, ICU discharge, or completion of the 28-day follow-up. | Same as the target trial. |
| **Causal Contrasts** | Per-protocol effect. | Same as the target trial. |
| **Analysis Plan** | To evaluate the effects of different transfusion strategies, it is necessary to account not only for baseline randomization but also for the differential risk arising from the fact that patients are managed under each transfusion strategy for varying lengths of time. | Clones were artificially censored at the time of deviation from the assigned strategy. To account for informative censoring and time-varying confounding, we applied stabilized inverse probability of censoring weights in a marginal structural model. We also used the parametric g-formula to estimate cumulative incidence functions while accounting for competing events (death and ICU discharge). |

### Supplementary Table 2. Eligibility Criteria, Exclusion Criteria, and Exposure Definitions

Time zero was defined as ICU admission (intime). Follow-up and all 24-hour intervals used for strategy assessment, covariate updating, and censoring were anchored to ICU admission. Where applicable, look-back windows for baseline covariates were defined relative to ICU admission or the suspected infection time, as specified below. Data were extracted from MIMIC-IV version 3.1, a de-identified electronic health record database from Beth Israel Deaconess Medical Center in Boston, Massachusetts, USA (2008–2023).

##### INCLUSION CRITERIA

| Variable | Codes / Item IDs | Data Sources and Operational Details |
| --- | --- | --- |
| **Age ≥18 years** | n/a | hosp_admissions and hosp_patients tables. Age was calculated as anchor_age + (year(admittime) − anchor_year) in accordance with the MIMIC-IV official age calculation method. Only patients aged ≥18 years at the time of hospital admission were eligible. |
| **Suspected infection: Microbiological culture** | All specimen types from hosp_microbiologyevents | MIMIC-IV hosp_microbiologyevents table, following the official suspicion_of_infection.sql implementation. All culture specimen types were included. A culture was considered eligible if the collection time fell between 24 hours before and 48 hours after ICU admission. |
| **Suspected infection: Antibiotic therapy** | Antibiotic drug names identified by text matching in hosp_prescriptions (see suspicion_of_infection.sql for the full list). Topical, ophthalmic, and otic routes were excluded. | MIMIC-IV hosp_prescriptions table. Antibiotic initiation was defined as the start of a new intravenous antibiotic prescription. The temporal relationship between culture collection and antibiotic initiation followed the official MIMIC-IV suspicion_of_infection.sql algorithm. The 4-day continuation rule was not applied to identify suspected infection at baseline. |
| **Suspected infection time** | Derived variable | Defined as the earlier of culture collection time and antibiotic start time among eligible events. The suspected infection time was required to occur within 72 hours of ICU admission. When multiple qualifying events were present, the earliest suspected infection time was used. |
| **Organ dysfunction: Cardiovascular (criterion a)** | Norepinephrine (Item ID 221906), Dopamine (221662), Epinephrine (221289), Phenylephrine (221749), Vasopressin (222315) | MIMIC-IV icu_inputevents table (sofa_cardiovascular.sql). Defined as the **new initiation** of any of the five specified vasopressor agents within the evaluation window. New initiation was identified as a transition from rate ≤0 (or no prior record) to rate >0 for a given agent, excluding continued infusions. Dobutamine (221653) and milrinone (221986) were extracted by the SQL query but explicitly excluded from the organ dysfunction criterion in the R implementation (cohort.R). |
| **Organ dysfunction: Respiratory (criterion b)** | Ventilator settings (Item IDs 224688, 224689, 224690, 224687, 224685, 224684, 224686, 224696, 220339, 224700, 223849, 229314, 223848, 224691), O₂ flow (223834, 227582, 227287), O₂ device (226732), FiO₂ (223835), SpO₂ (220277) | MIMIC-IV icu_chartevents table (sofa_respiratory.sql). Defined as the presence of any recorded ventilator setting, oxygen delivery device, or supplemental oxygen flow within the evaluation window. Records with O₂ device value of 'None' were excluded. The presence of any qualifying record was sufficient to meet this criterion; duration calculations were not required. |
| **Organ dysfunction: Renal (criterion c)** | Serum creatinine (Item ID 50912) | MIMIC-IV hosp_labevents table (sofa_labs.sql). Defined as serum creatinine ≥2.0× the baseline value, **or** estimated glomerular filtration rate (eGFR) decreased by ≥50% from the baseline eGFR. The eGFR was calculated using the CKD-EPI equation based on creatinine, age, sex, and race. Baseline creatinine was defined as the **minimum** creatinine value during the index hospitalization (hadm_id), and baseline eGFR as the **maximum** eGFR during the index hospitalization. Patients with **end-stage renal disease (ESRD)** were exempted from this criterion. ESRD was identified using ICD-9 code 5856 or ICD-10 code N186 from the hosp_diagnoses_icd table, with no time restriction (any time before or during the index admission). |
| **Organ dysfunction: Hepatic (criterion d)** | Total bilirubin (Item ID 50885) | MIMIC-IV hosp_labevents table. Defined as total bilirubin ≥2.0 mg/dL **and** ≥2.0× the baseline value. Baseline bilirubin was defined as the **minimum** total bilirubin during the index hospitalization. |
| **Organ dysfunction: Coagulation (criterion e)** | Platelet count (Item ID 51265) | MIMIC-IV hosp_labevents table. Defined as platelet count <100 ×10³/µL **and** ≤50% of the baseline value. This criterion was applied only when the baseline platelet count was ≥100 ×10³/µL. Baseline platelet count was defined as the **maximum** platelet count during the index hospitalization. |
| **Organ dysfunction: Metabolic (criterion f)** | Serum lactate (Item ID 50813) | MIMIC-IV hosp_labevents table. Defined as serum lactate ≥2.0 mmol/L within the evaluation window. No comparison with a baseline value was required. |
| **Organ dysfunction evaluation window** | n/a | All six organ dysfunction criteria (a–f) were evaluated within a time window of **±48 hours from the suspected infection time**, with an additional constraint that the event must occur within **72 hours of ICU admission** (i.e., win_end = min(suspected_infection_time + 48h, intime + 72h)). A patient was classified as having sepsis if **at least one** of the six criteria was met within this window. |
| **Baseline reference values** | n/a | For organ dysfunction criteria requiring comparison with baseline values (criteria c, d, and e), the baseline was defined as the least abnormal value observed during the index hospitalization (hadm_id): minimum for creatinine and bilirubin, maximum for platelet count and eGFR. This approach was adopted because pre-admission laboratory data were not available in MIMIC-IV, consistent with published recommendations. |

##### EXCLUSION CRITERIA

| Variable | Codes / Item IDs | Data Sources and Operational Details |
| --- | --- | --- |
| **ICU stay ≤48 hours** | n/a | MIMIC-IV icu_icustays table (exclusion_short_stay.sql). Defined as patients whose ICU length of stay, calculated as the difference between outtime and intime, was ≤48 hours (≤2,880 minutes). This criterion captured both patients who died and those who were discharged within 48 hours. These patients were excluded because the observation period was considered insufficient to differentiate transfusion strategies beyond the initial resuscitation phase. |
| **Massive hemorrhage** | **ICD-9**: 531, 5312, 5314, 5316 (gastric ulcer); 532, 5322, 5324, 5326 (duodenal ulcer); 533, 5332, 5334, 5336 (peptic ulcer, site unspecified); 534, 5342, 5344, 5346 (gastrojejunal ulcer); 53501, 5693, 578, 5781, 5789, 456, 4562 (other GI bleeding); 430 (subarachnoid hemorrhage); 431 (intracerebral hemorrhage); 432, 4321, 4329 (other intracranial hemorrhage); 853, 852, 8522, 8524 (traumatic intracranial injury); 280, 2851, 37272, 36281, 37923, 5118, 7191x, 5997, 5813, 5811, 580, 5812, 5804, 5818, 5819, 6262, 627, 6268, 6269, 6271, 7847, 7848, 7863, 56881, 38861, 78499 (other bleeding). **ICD-10**: K250, K252, K254, K256; K260, K262, K264, K266; K270, K272, K274, K276; K280, K282, K284, K286; K290, K625, K920, K921, K922, I850, I983; I60, I600–I609; I61, I610–I619; I62, I620, I621, I629; S063–S066; D500, D62, H113, H356, H431, H450, J942, M250, N02, N020–N029, N920, N921, N924, N938, N939, N950, R04, R040–R042, R048, R049, R31, K661, H922, R58. | MIMIC-IV hosp_diagnoses_icd table (exclusion_massive_hemorrhage.sql). Defined as any primary or secondary ICD-9 or ICD-10 diagnosis code recorded during the **index hospital admission** (hadm_id). Patients were excluded at the admission level—i.e., only the current admission’s diagnosis codes were assessed (not lifetime history). ICD codes in MIMIC-IV are stored without decimal points. Code selection was based on published exclusion criteria for transfusion studies. |
| **Hematologic diseases** | **ICD-9**: 201–208 (malignant neoplasms of lymphoid/hematopoietic tissue); 280–289 (diseases of blood and blood-forming organs); 279 (disorders of immune mechanism); 135 (sarcoidosis). **ICD-10**: C81–C96 (malignant neoplasms); D50–D77 (diseases of blood); D80–D84, D86, D89 (immune disorders). | MIMIC-IV hosp_diagnoses_icd table (exclusion_hematologic.sql). Defined using **prefix matching** (e.g., LIKE '201%') to capture all subcodes within each category. Codes were assessed across **all hospital admissions** for each patient (subject_id), reflecting a lifetime comorbidity rather than current-admission-only exclusion. Patients were excluded because their transfusion management is driven by hematologic indications fundamentally different from sepsis-related anemia. |
| **Comfort measures only (CMO)** | CMO status: icu_chartevents Item ID 223758, value = 'Comfort measures only'. Hemoglobin: hosp_labevents Item ID 51222. | MIMIC-IV icu_chartevents and hosp_labevents tables (exclusion_cmo.sql). Patients were excluded if their **first Hgb measurement time** was strictly **after** their **first documented CMO decision time** within the same admission. This implemented the protocol criterion of excluding patients “for whom a CMO care goal had been established before any Hgb monitoring occurred.” The CMO documentation source was icu_chartevents (Item ID 223758), consistent with the MIMIC-IV official code_status.sql concept definition. The poe (Physician Order Entry) table was not used, as it does not typically contain CMO orders in MIMIC-IV. |

##### ADDITIONAL COHORT RESTRICTIONS

| Variable | Codes / Item IDs | Data Sources and Operational Details |
| --- | --- | --- |
| **Readmission within 48 hours** | n/a | MIMIC-IV icu_icustays table (implemented in cohort.R). For the same patient (subject_id), consecutive ICU stays were sorted by intime. If the time interval between the previous stay’s outtime and the current stay’s intime was **<48 hours**, the subsequent stay was excluded. This restriction avoided non-independent observations from closely spaced ICU episodes that likely represent a single clinical course. |
| **Limited hemoglobin measurement** | Hemoglobin: Item IDs 50811 (blood gas) and 51222 (complete blood count) from hosp_labevents | MIMIC-IV hosp_labevents and icu_icustays tables (flag_hgb_limited_measurement.sql). ICU stay duration was divided into 24-hour intervals (“buckets”) starting from ICU admission (intime). Bucket 0 corresponded to the first 24 hours; the discharge bucket was the interval containing outtime. Patients were **flagged** (is_limited_hgb = 1) if all Hgb measurements occurred exclusively in Bucket 0 and/or the discharge bucket, with **no measurements in any intermediate bucket**. These patients were **retained** in the primary analysis. |

##### EXPOSURE DEFINITION

| Variable | Codes / Item IDs | Data Sources and Operational Details |
| --- | --- | --- |
| **Red blood cell (RBC) transfusion events** | Item IDs: 220996 (Packed Red Blood Cells), 225168 (Packed RBC) from icu_inputevents | MIMIC-IV icu_inputevents table (transfusion_rbc.sql). Each RBC transfusion event was identified by the presence of an input event record with a qualifying Item ID. The start time (starttime) of the transfusion was used to assign transfusion events to 24-hour intervals for daily strategy assessment. |
| **Daily hemoglobin for strategy assignment** | Hemoglobin: Item IDs 50811 and 51222 from hosp_labevents | MIMIC-IV hosp_labevents table (implemented in 5_longitudinal_data.R). For each 24-hour interval (day), the Hgb value used for strategy assignment was defined conditionally: (1) **on days with transfusion**, the mean of Hgb values measured **before** the first transfusion start time of that day was used (approximating the trigger Hgb that prompted the transfusion decision); (2) **on days without transfusion**, the mean of all Hgb values measured during that day was used. |
| **Transfusion strategy classification** | n/a | Each patient-day was classified according to one of three transfusion strategies based on the daily Hgb value and transfusion status: (1) restrictive strategy, in which RBC transfusion was permitted only when Hgb was ≤7.0 g/dL; (2) liberal strategy, in which RBC transfusion was permitted when Hgb was >7.0 g/dL; and (3) no-transfusion strategy, in which no RBC transfusion was permitted. Strategy adherence was assessed in consecutive 24-hour intervals from ICU admission (Day 0). A patient-clone was artificially censored at the first deviation from the assigned strategy. |
| **Follow-up period** | n/a | We began follow-up at Day 0 (ICU admission) and continued it for up to 28 days. Follow-up was terminated at the earliest occurrence of: (1) deviation from the assigned transfusion strategy (artificial censoring), (2) ICU-acquired infection (primary outcome), (3) death, (4) ICU discharge, or (5) completion of the 28-day observation period. |

*Abbreviations: CKD-EPI, Chronic Kidney Disease Epidemiology Collaboration; CMO, comfort measures only; eGFR, estimated glomerular filtration rate; ESRD, end-stage renal disease; FiO₂, fraction of inspired oxygen; GI, gastrointestinal; Hgb, hemoglobin; ICD, International Classification of Diseases; ICU, intensive care unit; IV, intravenous; MIMIC-IV, Medical Information Mart for Intensive Care IV; RBC, red blood cell; SOFA, Sequential Organ Failure Assessment; SpO₂, peripheral oxygen saturation.*

### Supplementary Table 3. Operational Definition of ICU-Acquired Infections and Competing Events

ICU-acquired infection was defined using a prespecified algorithm combining microbiological and antimicrobial data. The rules below describe the implementation used to construct the primary outcome.

| Criterion | Operational Rule | Data Source & Detailed Implementation |
| --- | --- | --- |
| **1. Microbiological Criteria** | A qualifying blood culture obtained during the ICU stay. | MIMIC-IV hosp_microbiologyevents table (outcome_infection.sql). Only records with a specimen type containing “BLOOD CULTURE” were included. The culture time (charttime or chartdate) was required to fall between the ICU admission (intime) and ICU discharge (outtime). |
| **2. Antibiotic Criteria** | Initiation of a new intravenous (IV) antibiotic within ±48 hours of the qualifying blood culture. | MIMIC-IV hosp_prescriptions table (outcome_infection.sql, active_antibiotics.sql). Antibiotics were restricted to those administered via the IV route (route starting with “IV” or “INTRAVEN”). A new start was defined as initiation of a specific intravenous antibiotic with no algorithmically linked administration of the different antibiotic in the preceding 24 hours. The new start time was required to be ≥ culture_time - 48h and ≤ culture_time + 48h. |
| **3. Four-Day Rule (Duration)** | The new antibiotic regimen must be administered continuously for at least **4 days (96 hours)**. | Computed in R (0_utils.R, check_4day_rule). A course was considered continuous if the gap between consecutive administrations of any qualifying IV antibiotics was **≤24 hours**. The continuous sequence must cover the period from the initial start time ($S$) to $S+96$ hours. |
| **4. Exceptions to the Four-Day Rule** | The 96-hour duration requirement was modified for patients who experienced a competing event (death, ICU discharge, or hospital transfer) prior to completing the full course. | Computed in R (0_utils.R). If the earliest competing event occurred before the standard 96-hour completion time, the requirement was considered met if the antibiotic course was continued until **at least 24 hours prior to the event** (required_end = max(start_time, event_time - 24 hours)). |
| **5. Timing of Infection Onset** | The onset time was defined as the earlier of the blood culture time or the new IV antibiotic start time. | SQL logic: LEAST(culture_time, antibiotic_time). This time point was taken to represent the onset of suspected infection. |
| **6. ICU-Acquired Restriction** | The suspected infection must occur strictly **>72 hours after ICU admission**. | SQL logic (outcome_infection.sql): culture_time > intime + 72 hours. Any infection meeting the above criteria but occurring ≤72 hours after ICU admission was considered present-on-admission rather than ICU-acquired and was not counted as an outcome event. |
| **7. Competing event: Death** | In-hospital death during the ICU stay. | deathtime from MIMIC-IV hosp_admissions table. When deathtime was unavailable, the hospital discharge time (dischtime) was used for patients with hospital_expire_flag = 1. Implemented in 4_outcomes.R. |
| **8. Competing event: ICU discharge** | Discharge from the ICU alive. | Defined as the recorded ICU departure time (outtime) from icu_icustays. |

### Supplementary Figure 1. Directed Acyclic Graphs (DAGs) for Confounder Selection

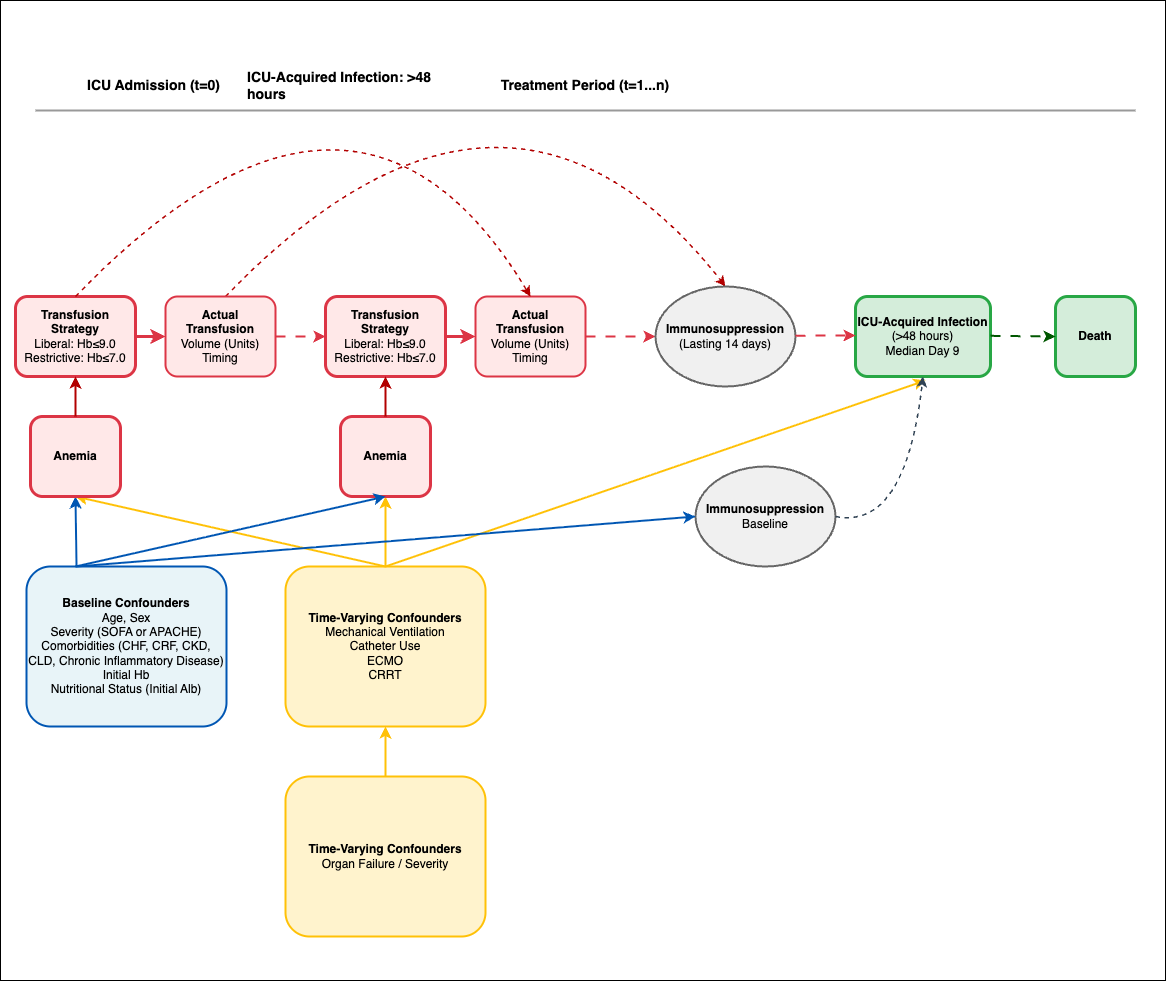

*Directed Acyclic Graph (DAG) for Transfusion and Infection*

### Supplementary Table 4. Covariate Definitions

The following baseline and time-varying covariates were included in the inverse probability of censoring weight (IPCW) models. Baseline covariates were measured before or at time zero, whereas time-varying covariates were updated daily during follow-up.

| Variable | Codes / Item IDs | Data Sources and Operational Details |
| --- | --- | --- |
| Age | hosp_patients.anchor_age | Age at admission, extracted from hosp_patients. |
| Baseline SOFA score | Respiration: 50821 (PaO₂), 50816 (FiO₂ lab), 223835 (FiO₂ chart), 220277 (SpO₂);  Coagulation: 51265 (Platelets);  Liver: 50885 (Bilirubin);  Cardiovascular: 220052, 220181, 225312 (MAP); 221906 (Norepinephrine), 221662 (Dopamine), 221289 (Epinephrine), 221653 (Dobutamine);  CNS: 220739 (GCS Eye), 223900 (GCS Verbal), 223901 (GCS Motor);  Renal: 50912 (Creatinine);  Urine output: 226559, 226560, 226561, 226563, 226564, 226565, 226567, 226584, 227510, 227519 | Calculated as the total score (0–24) on Day 0 of the longitudinal dataset, derived from 6 organ components (see footnote below). Computed using the worst values recorded during the first 24 hours following ICU admission (or missing if completely unobserved). |
| Baseline hemoglobin | Item IDs: 50811, 51222 | Defined as the measurement closest to ICU admission, within a window of ±24 hours (hosp_labevents). |
| Serum albumin | Item ID: 50862 | Defined as the measurement closest to ICU admission, within a window of ±24 hours (hosp_labevents). |
| Body Mass Index (BMI) | Weight: 226512, 226531;  Height: 226730, 226707 | Calculated as Weight (kg) / Height (m)², using the weight closest to ICU admission and the median recorded height (icu_chartevents). |
| Congestive heart failure | Defined by Charlson Comorbidity Index criteria | Derived from hosp_diagnoses_icd using definitions adapted from the official MIMIC-IV charlson.sql concept script. |
| Chronic pulmonary disease | Defined by Charlson Comorbidity Index criteria | Derived from hosp_diagnoses_icd using definitions adapted from the official MIMIC-IV charlson.sql concept script. |
| Renal disease | Defined by Charlson Comorbidity Index criteria | Derived from hosp_diagnoses_icd using definitions adapted from the official MIMIC-IV charlson.sql concept script. |
| Liver disease (mild or severe) | Defined by Charlson Comorbidity Index criteria | Composite of mild and severe liver disease, derived from hosp_diagnoses_icd using definitions adapted from the official MIMIC-IV charlson.sql concept script. |
| Rheumatic disease | Defined by Charlson Comorbidity Index criteria | Derived from hosp_diagnoses_icd using definitions adapted from the official MIMIC-IV charlson.sql concept script. |
| Daily hemoglobin | Item IDs: 50811, 51222 | On days with transfusion, daily Hgb was defined as the mean of measurements obtained before the first transfusion event of that day, to approximate the pre-transfusion trigger value. On days without transfusion, daily Hgb was defined as the mean of all measurements obtained that day. Treated as missing if no measurements were available (hosp_labevents). |
| Daily SOFA score | Same as Baseline SOFA | Maximum (worst) SOFA score over each 24-hour observation window (Day 1 to 27), calculated using the same 6-component algorithm as the baseline SOFA score. |

*SOFA score calculation.* The total SOFA score (range 0–24) was computed as the sum of six organ-specific component scores, each ranging from 0 to 4: (1) **Respiration** — based on the PaO₂/FiO₂ ratio; when PaO₂ was unavailable, the SpO₂/FiO₂ ratio was used as a surrogate (S/F pathway); patients with SpO₂ ≥ 98% and no PaO₂/FiO₂ data were assumed to have normal respiratory function (score 0); (2) **Coagulation** — based on platelet count; (3) **Liver** — based on total bilirubin; (4) **Cardiovascular** — based on mean arterial pressure (MAP) and vasopressor infusion rates (norepinephrine, dopamine, epinephrine, dobutamine); when MAP was unavailable, the vasopressor dose alone determined the score; (5) **Central nervous system** — based on the Glasgow Coma Scale (GCS) total score; (6) **Renal** — based on serum creatinine; when creatinine was unavailable, daily urine output was used as a fallback. For each component, the worst (highest) value within each 24-hour observation window was used. All SOFA definitions were adapted from the official MIMIC-IV concept scripts (sofa.sql).

### Supplementary Table 5. Baseline Characteristics and Missingness at Study Baseline (N = 4,013)

| Variable | N Valid | Summary statistic, mean (SD) / median [IQR] / n (%) | Missing, n (%) |
| --- | --- | --- | --- |
| **Demographics** |  |  |  |
| Age (years) | 4013 | 64.5 (17.7) / 67.0 [54.0-78.0] | 0 (0.00%) |
| Female sex | 4013 | 1657 (41.3%) | 0 (0.00%) |
| BMI (kg/m²) | 2837 | 29.6 (8.2) / 28.0 [24.3-33.1] | 1176 (29.30%) |
| Weight (kg) | 2892 | 84.6 (25.1) / 80.3 [68.0-96.5] | 1121 (27.93%) |
| **Severity and Physiology** |  |  |  |
| Baseline SOFA score | 1781 | 5.3 (3.2) / 5.0 [3.0-8.0] | 2232 (55.62%) |
| Mean arterial pressure (mmHg) | 3989 | 81.0 (10.6) / 79.0 [73.5-87.2] | 24 (0.60%) |
| GCS | 4003 | 8.0 (4.7) / 7.0 [3.0-13.5] | 10 (0.25%) |
| Oxygenation (P/F or S/F ratio) | 3070 | 137.6 (97.9) / 105.0 [75.0-176.0] | 943 (23.50%) |
| **Laboratory Values** |  |  |  |
| Baseline hemoglobin (g/dL) | 4008 | 11.9 (2.2) / 11.9 [10.3-13.4] | 5 (0.12%) |
| Creatinine (mg/dL) | 3980 | 1.2 (1.0) / 1.0 [0.7-1.3] | 33 (0.82%) |
| Bilirubin (mg/dL) | 1790 | 1.2 (2.4) / 0.6 [0.4-1.1] | 2223 (55.39%) |
| Platelets (x10³ / µL) | 3970 | 212.7 (98.3) / 194.0 [146.0-254.8] | 43 (1.07%) |
| Albumin (g/dL) | 1247 | 3.2 (0.6) / 3.2 [2.7-3.6] | 2766 (68.93%) |
| **Life Support and Comorbidities** |  |  |  |
| Mechanical ventilation | 4013 | 2725 (67.9%) | 0 (0.00%) |
| Vasopressor use | 4013 | 1014 (25.3%) | 0 (0.00%) |
| Congestive heart failure | 4013 | 1115 (27.8%) | 0 (0.00%) |
| Chronic pulmonary disease | 4013 | 1114 (27.8%) | 0 (0.00%) |
| Renal disease | 4013 | 507 (12.6%) | 0 (0.00%) |
| Liver disease | 4013 | 264 (6.6%) | 0 (0.00%) |
| Rheumatic disease | 4013 | 114 (2.8%) | 0 (0.00%) |

### Supplementary Figure 2. MICE Imputation Quality Audit: Distributional Comparison of Observed vs. Imputed-Only Values

A. **Daily Hemoglobin**

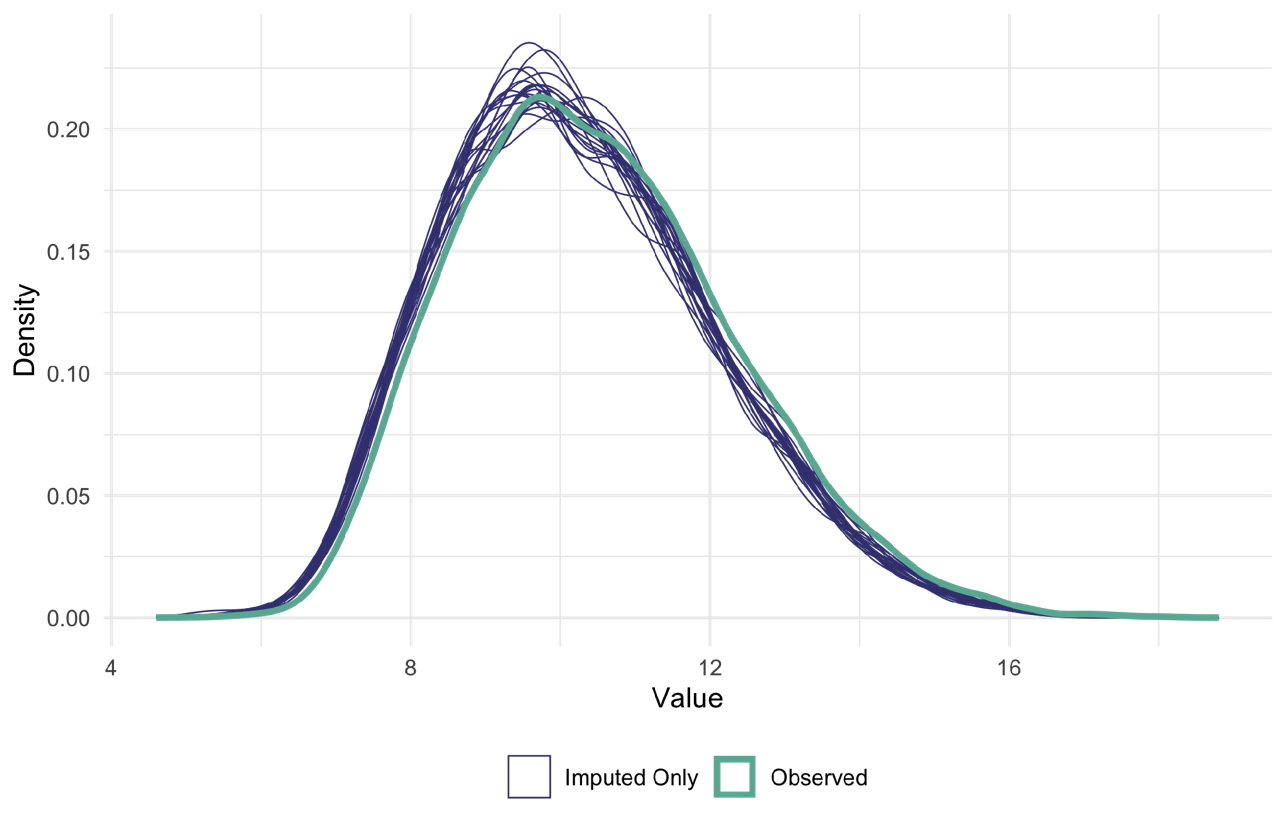

B. **Platelets**

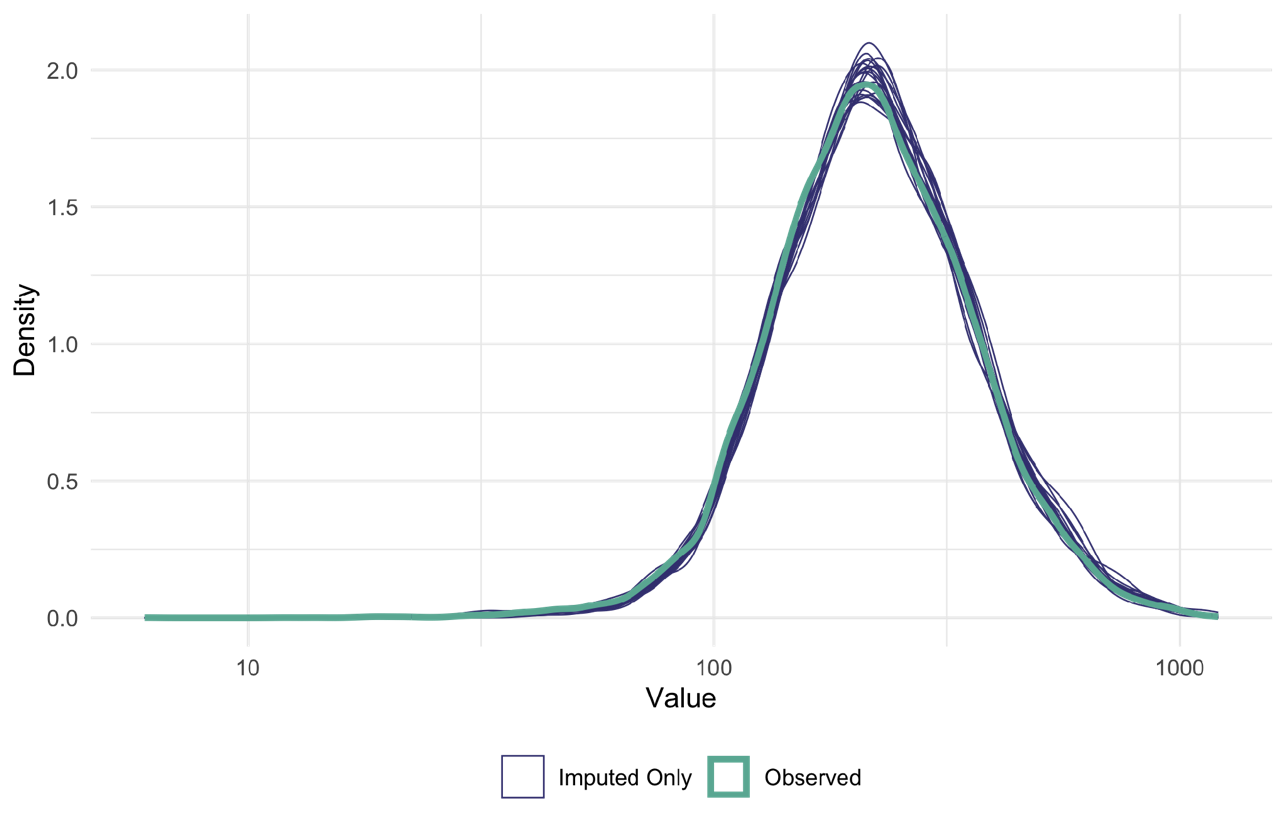

C. **Bilirubin**

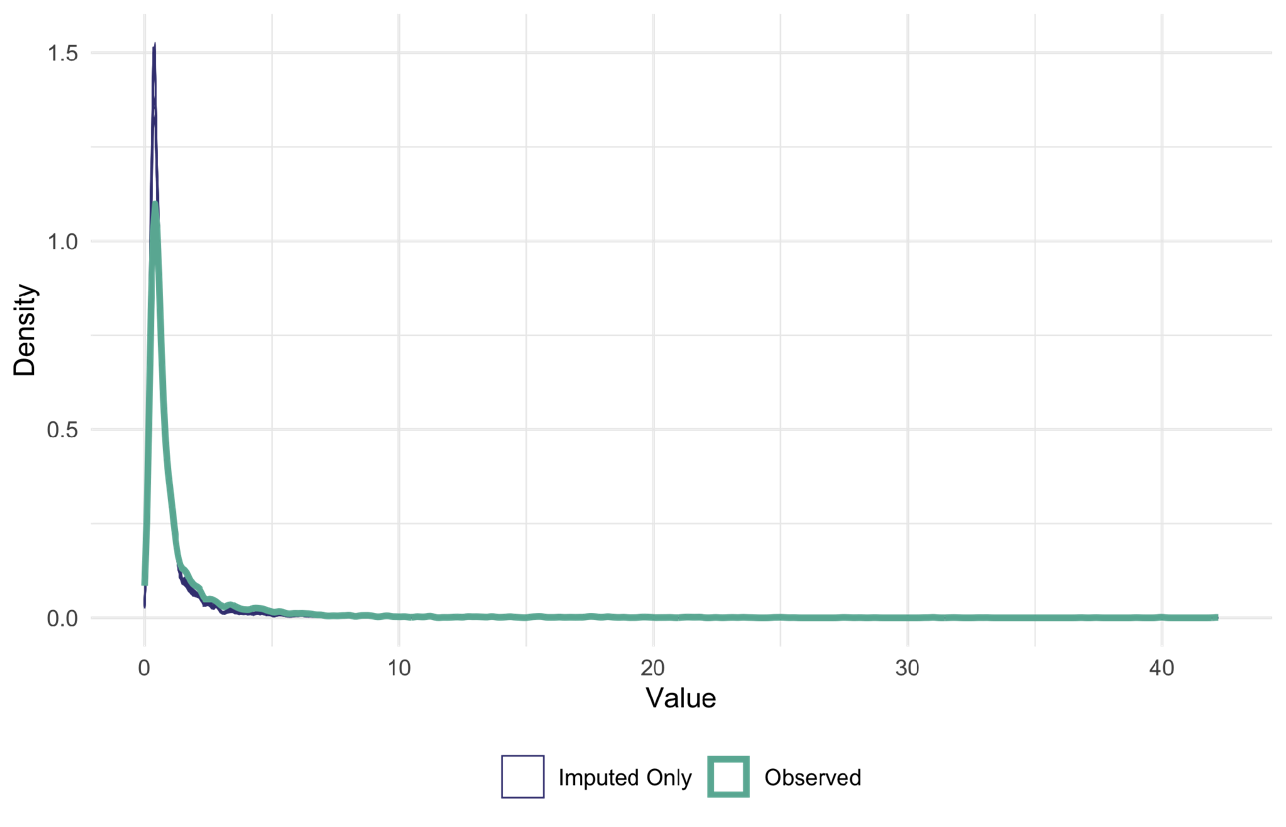

D. **Creatinine**

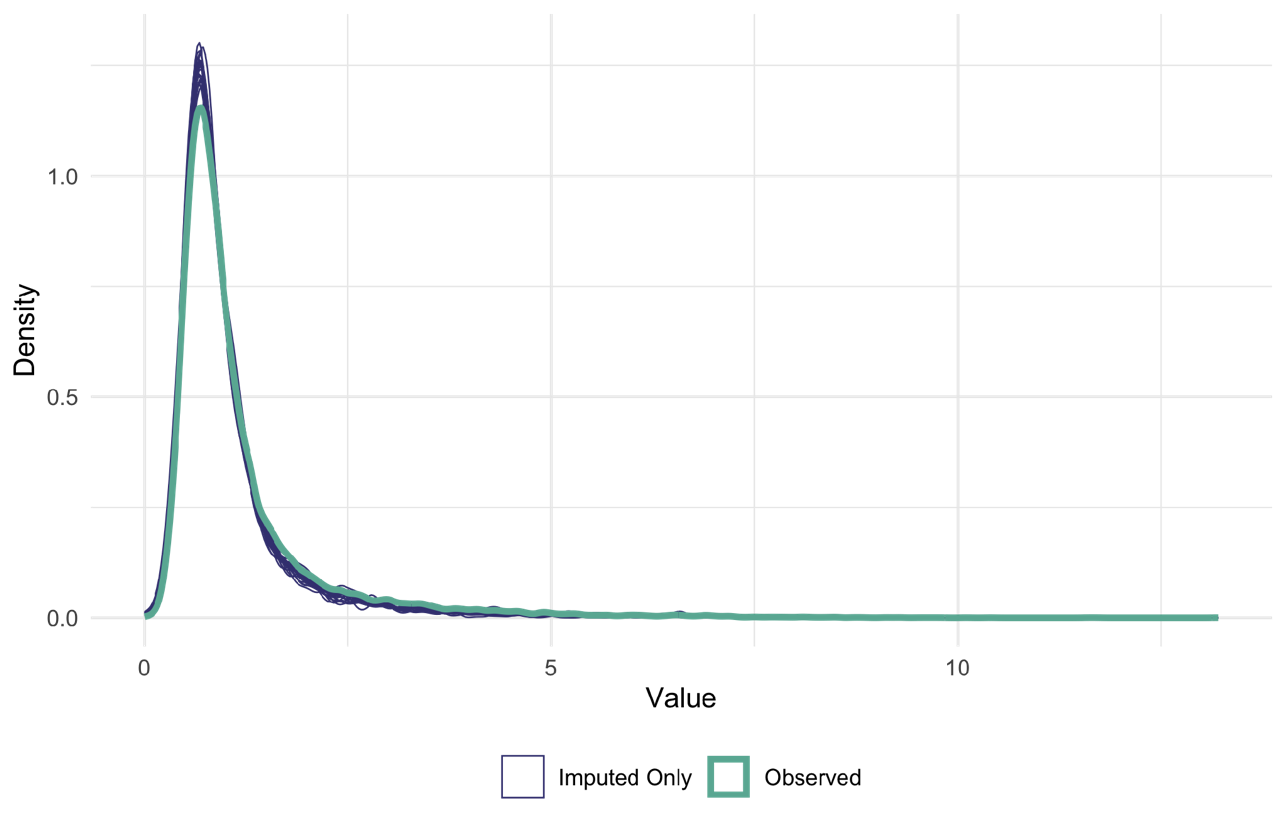

E. **Daily Urine Output**

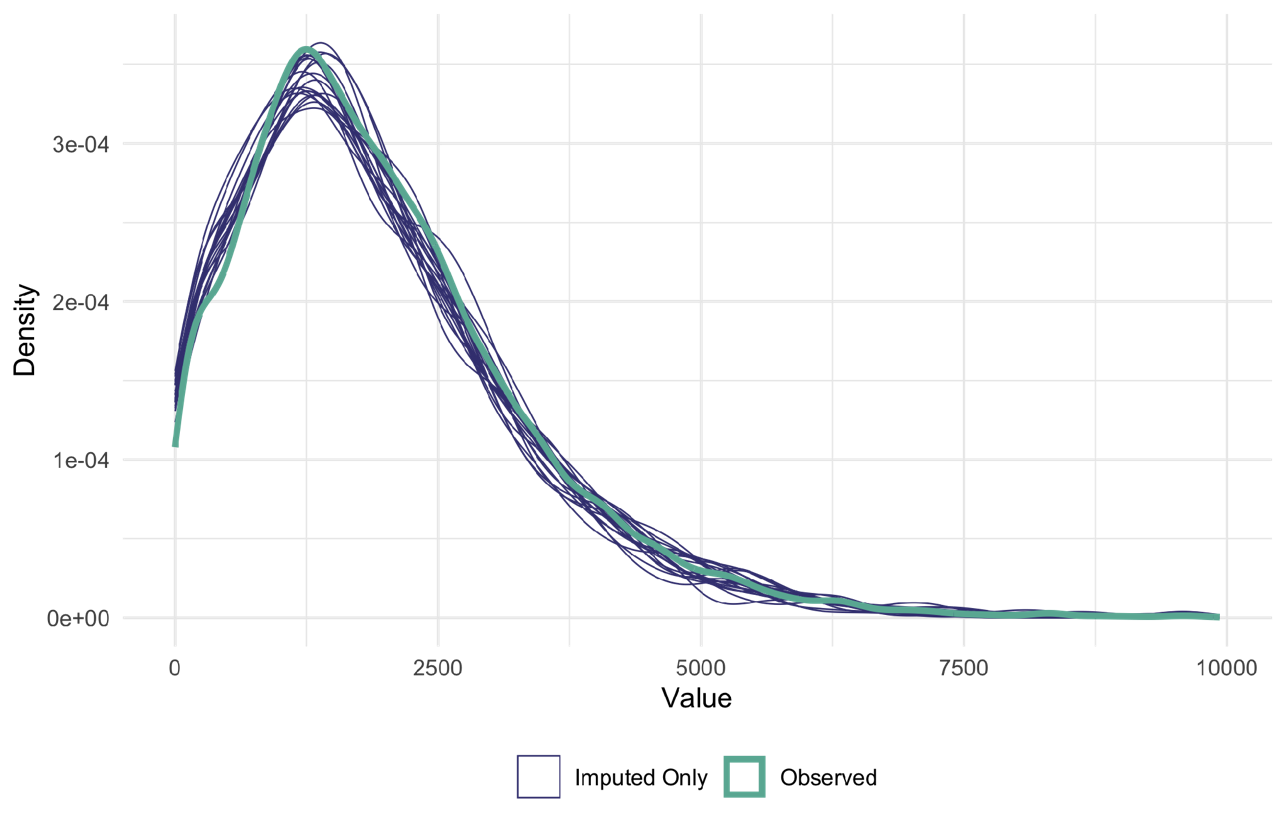

F. **Glasgow Coma Scale (GCS)**

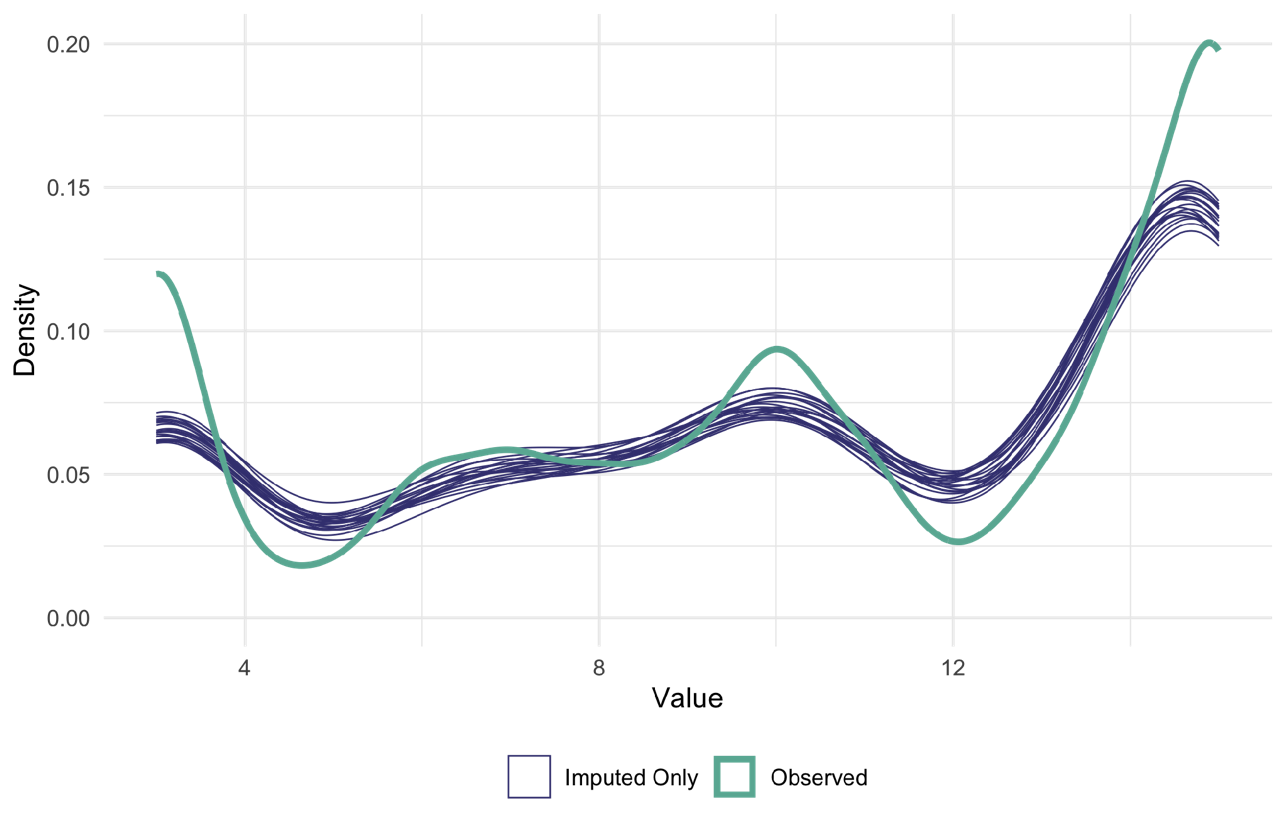

**G. PaO₂**

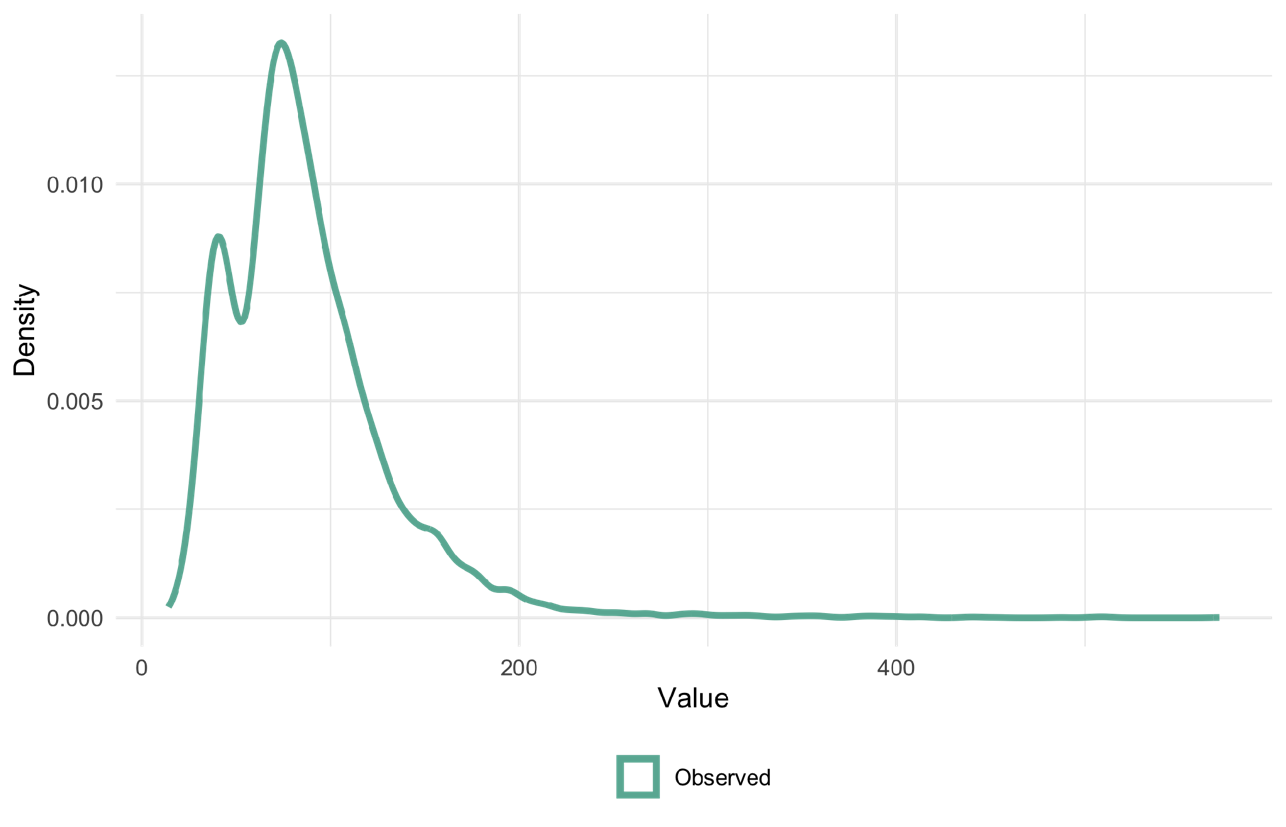

**H. SpO₂**

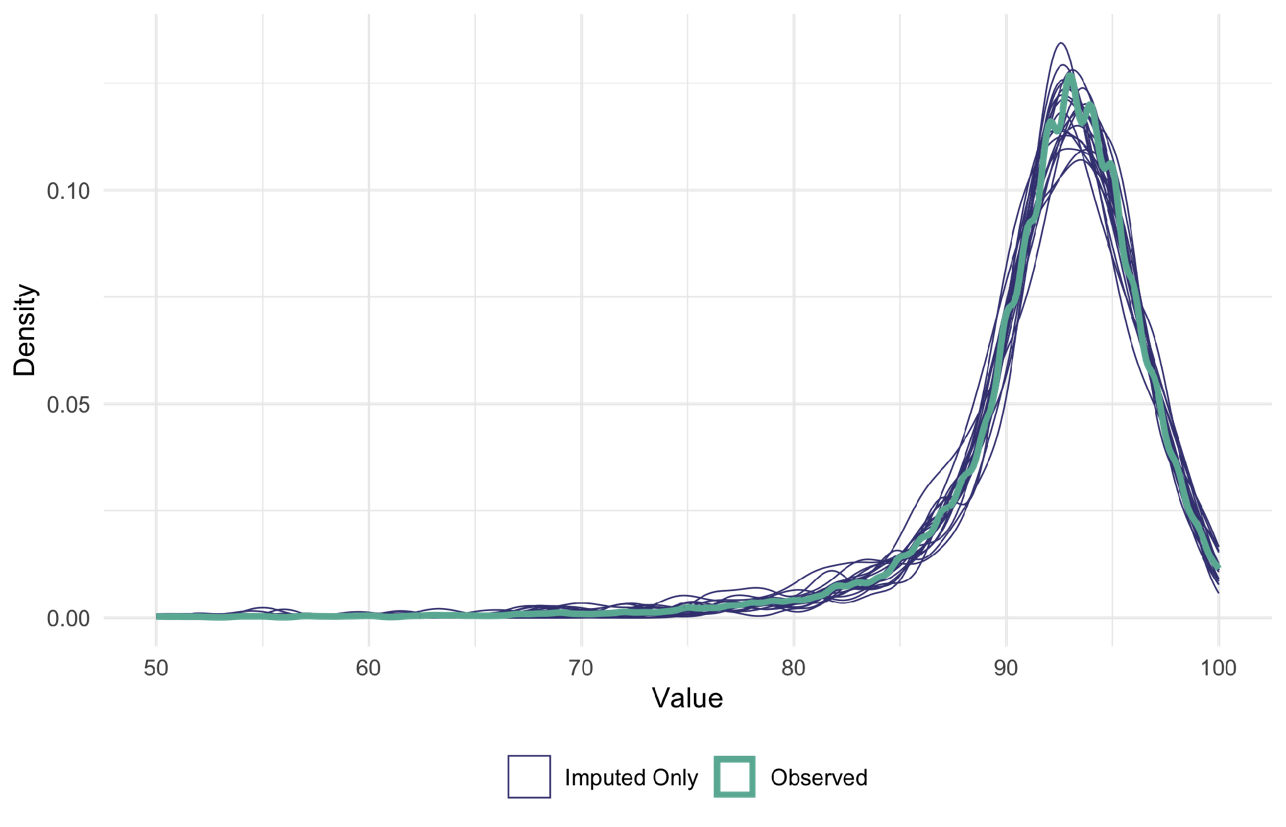

**I. FiO₂**

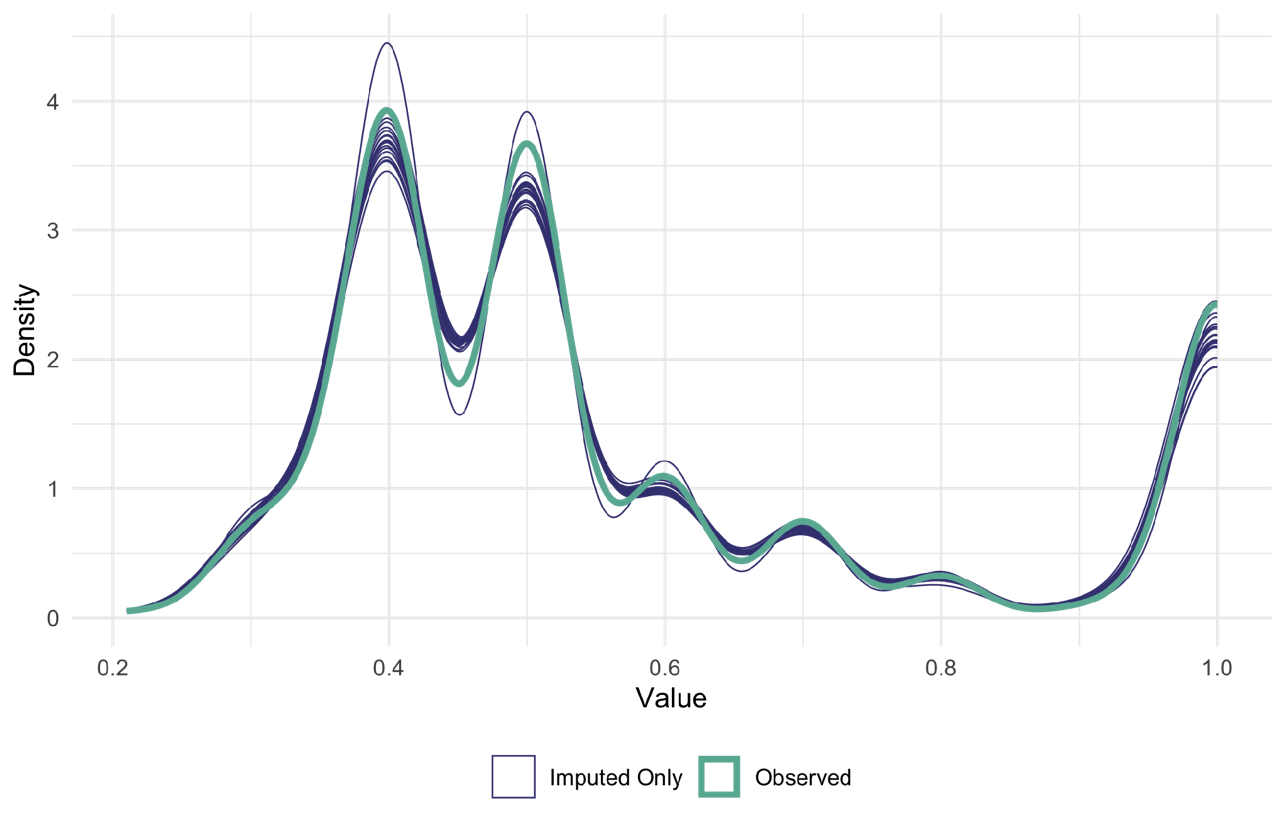

**J. Systolic Blood Pressure**

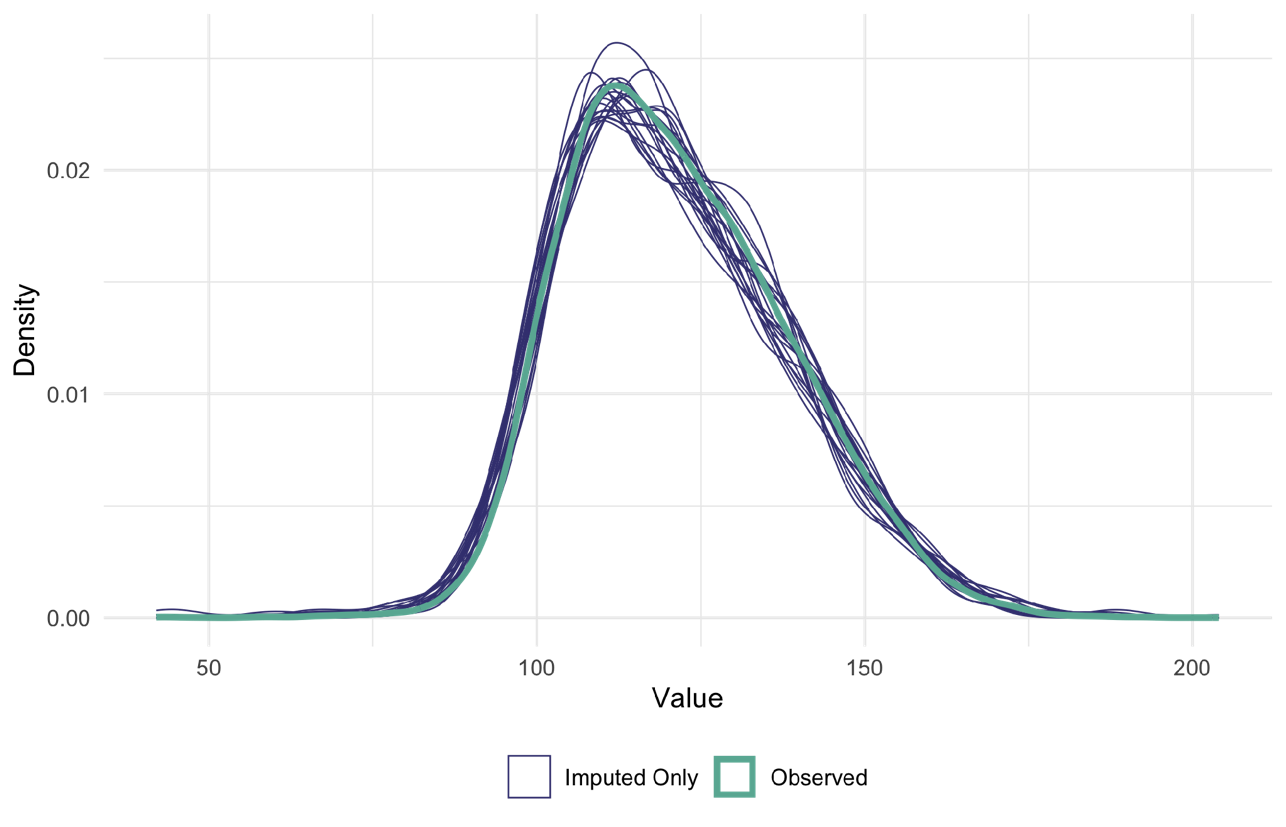

**K. Diastolic Blood Pressure**

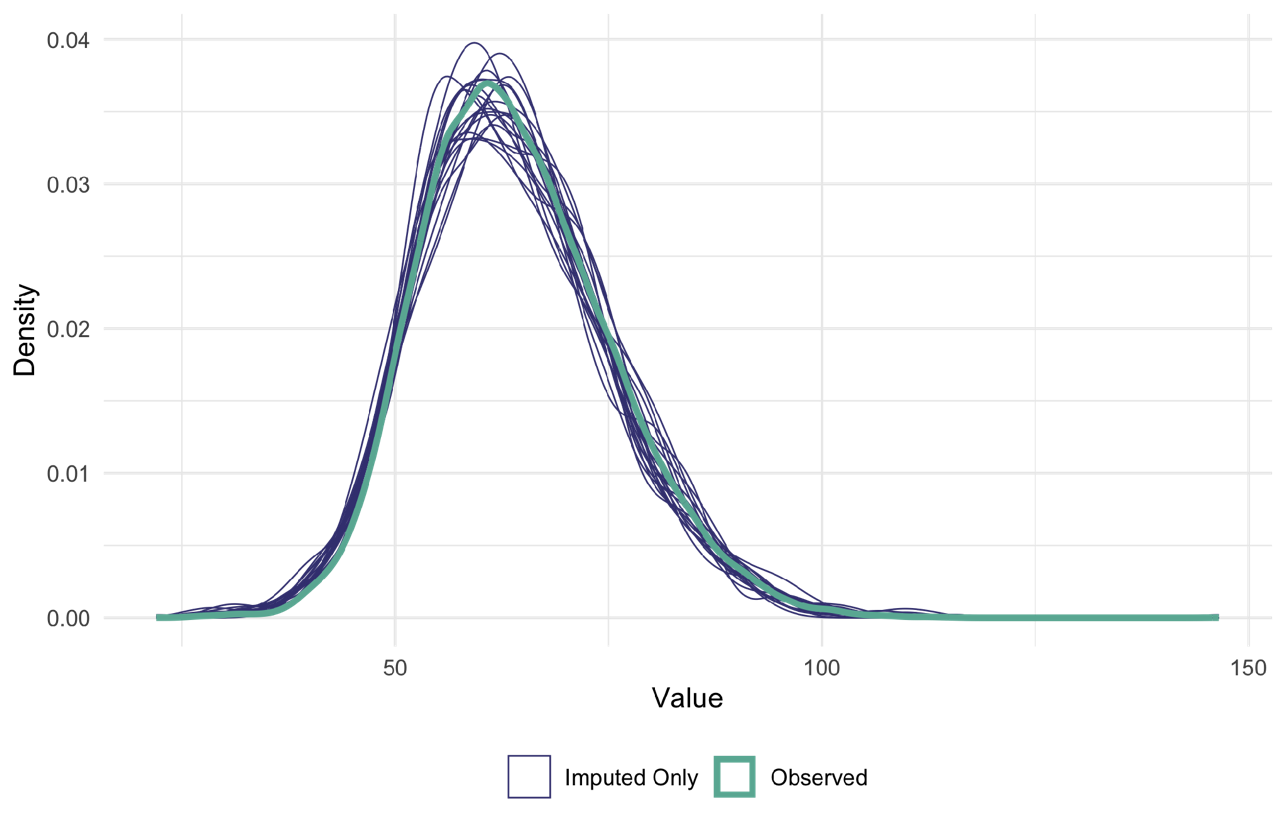

**L. Baseline SOFA Score**

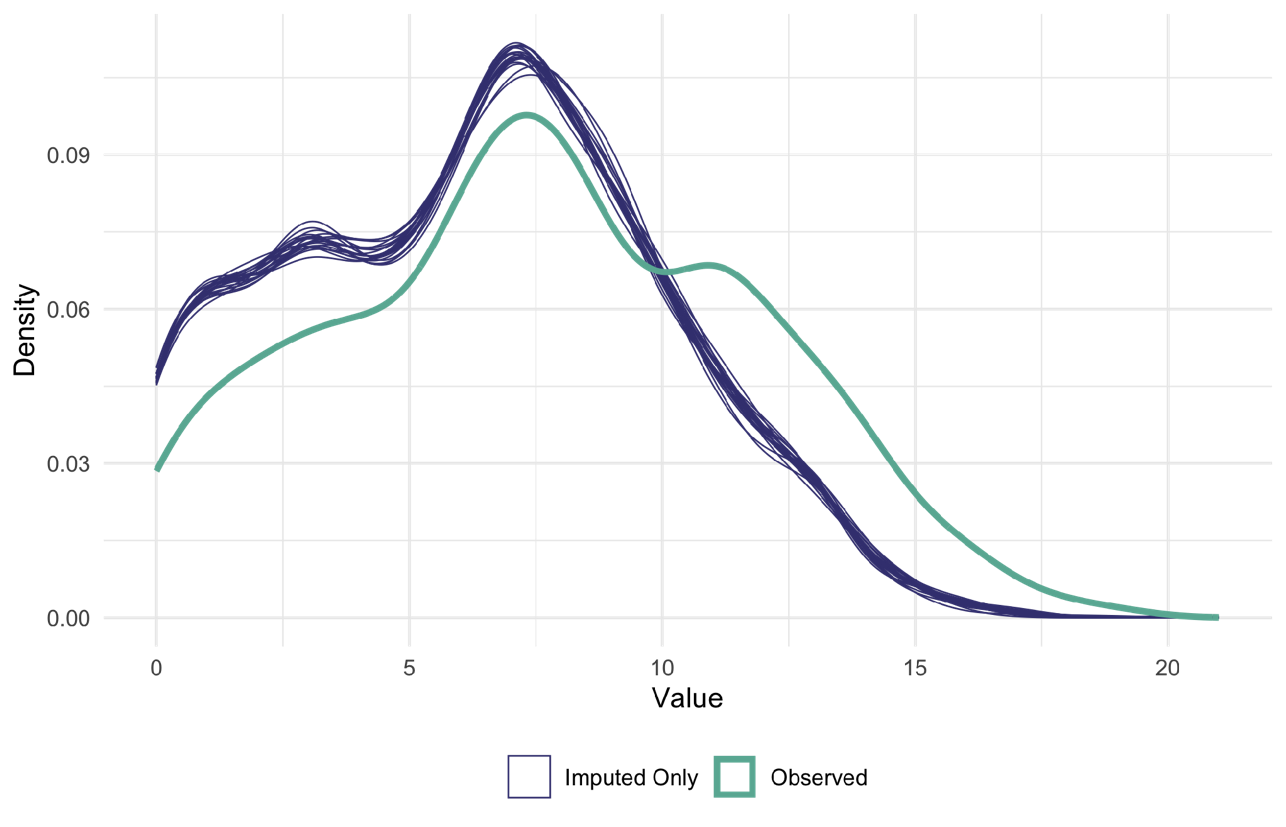

*Figure note: Kernel density plots compare the distributions of observed values (blue) and imputed-only values (red) across 20 imputed datasets. The broadly similar distributions support the plausibility of the imputation model under the missing-at-random assumption.*

### Supplementary Methods for Statistical Analysis

To emulate transfusion strategies defined by Hgb thresholds, we specified a hypothetical target trial in which patients with sepsis were assigned at time zero to one of three strategies: a restrictive strategy (transfusion permitted when Hgb was ≤7.0 g/dL), a liberal strategy (transfusion permitted when Hgb was >7.0 g/dL), or a no-transfusion strategy. Because transfusion practice was not protocolized in the observational data, observed treatment patterns were heterogeneous relative to the prior randomized trial. We therefore implemented the observed data using these three prespecified strategies.

##### Primary Estimand

The primary estimand was the per-protocol effect of each transfusion strategy on the 28-day risk of ICU-acquired infection. We estimated this effect using a marginal structural model fitted to cloned observational data with inverse probability of censoring weights. Effect estimates are presented as odds ratios (ORs) with 95% confidence intervals (CIs). Adherence to the assigned strategy was modeled over time using logistic regression conditional on time-varying covariates and baseline covariates, and inverse probability of censoring weights (IPCW) were applied to adjust for informative censoring due to deviations from the assigned strategy. The OR was interpreted as the per-protocol effect while patients remained alive and under observation in the ICU during the 28-day follow-up.

##### Clone-Censor-Weight Approach

To emulate the per-protocol effect of the target trial, we adopted the clone-censor-weight approach. At time zero, each eligible patient was cloned into three copies, with each clone assigned to one of the three transfusion strategies. Strategy adherence was assessed in 24-hour intervals for 28 days, and a clone was artificially censored at the first deviation from its assigned strategy. Follow-up ended at the earliest occurrence of artificial censoring, ICU-acquired infection, death, ICU discharge, or Day 28.

At time zero, each eligible patient was cloned into three copies, one for each strategy. Baseline covariates measured before or at the start of follow-up included age, baseline SOFA score, baseline Hgb, albumin, body mass index (BMI), congestive heart failure, chronic pulmonary disease, renal disease, liver disease, and rheumatic disease.

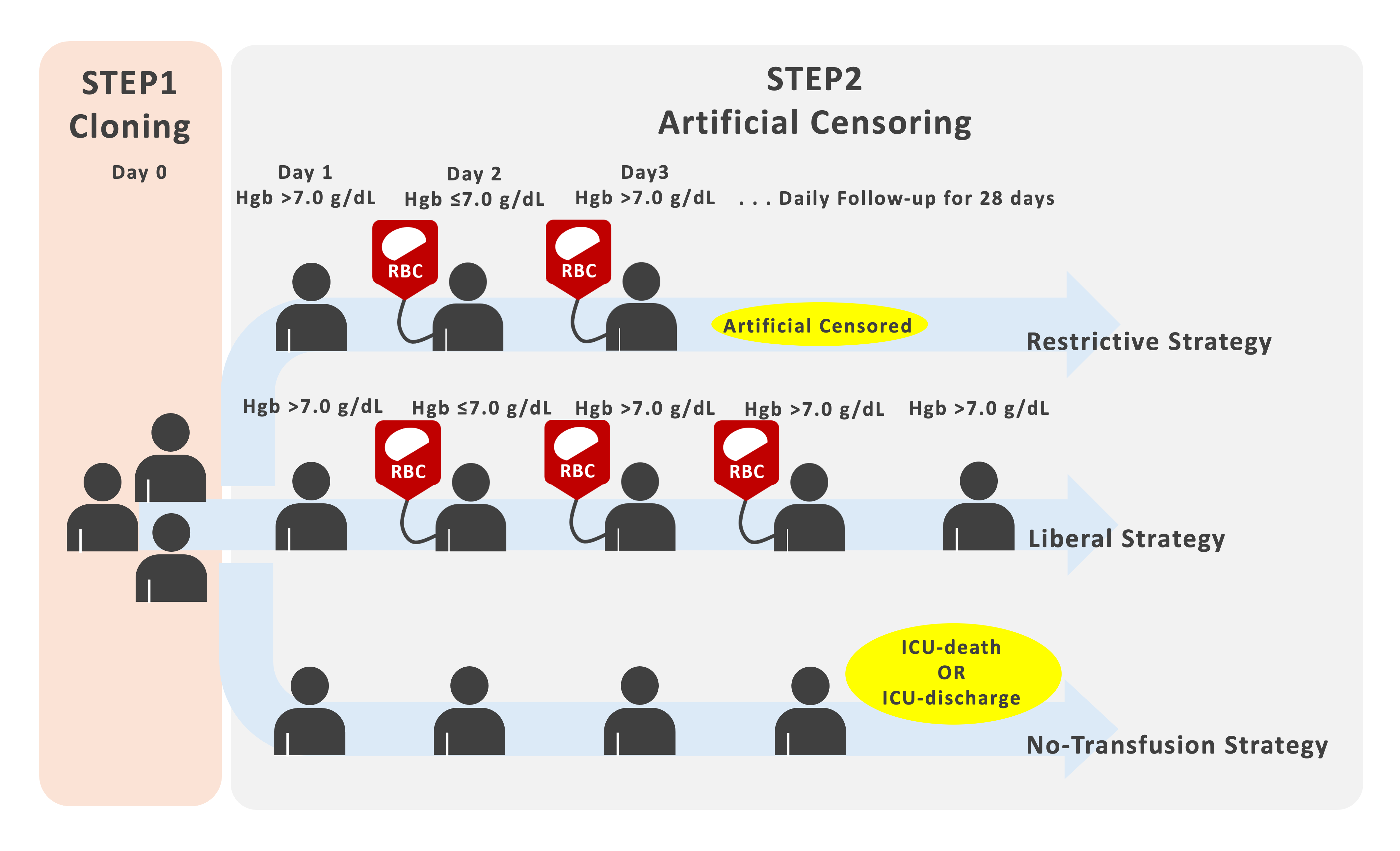

##### Marginal Structural Model

We estimated the per-protocol effect using a marginal structural model implemented as weighted pooled logistic regression. Stabilized inverse probability of censoring weights were estimated daily from pooled logistic models for adherence to the assigned strategy. The denominator model included assigned strategy, time, baseline covariates, and daily time-varying covariates, whereas the numerator model included assigned strategy, time, and baseline covariates only. Time was modeled using natural cubic splines with 2 degrees of freedom. To improve numerical stability, predicted probabilities were bounded between 0.01 and 0.99, and cumulative stabilized weights were truncated at the 97.5th percentile. Standard errors were estimated using cluster-robust sandwich estimators at the patient level. Analyses were conducted across 20 multiply imputed datasets, and estimates were combined using Rubin’s rules.

##### Secondary Analysis: Parametric g-formula

We estimated the 28-day cumulative incidence of ICU-acquired infection under each strategy using the parametric g-formula. We specified parametric models for daily Hgb, daily SOFA score, daily transfusion status, and the competing event process. Outcomes were modeled in discrete time as four mutually exclusive states: ICU-acquired infection, ICU discharge, in-ICU death, and no event. All models were conditioned on baseline covariates and covariate history, including first-order lags of time-varying covariates and prior transfusion status. Time since ICU admission was modeled using natural cubic splines (4 degrees of freedom), and admission year was included as a continuous term. To reduce the risk of perfect separation due to sparse events, infrequent ICU categories with zero events were collapsed into an “Other” category.

We then performed Monte Carlo simulation to generate the joint evolution of covariates, transfusion status, and outcomes over 28 days under each strategy. At Day 0, covariates were initialized to their observed baseline values. On each subsequent day, transition probabilities were estimated using a multinomial logit parameterization so that the probabilities across the four states summed to 1. Simulated trajectories were updated sequentially by drawing covariates, transfusion status, and outcomes forward one day at a time.

Cumulative incidence functions were estimated as the proportion of simulated individuals experiencing each event by each day, based on the corresponding cause-specific hazards. Risk differences were calculated as differences in infection CIFs between strategies over the 28-day follow-up. Uncertainty was quantified using a nonparametric bootstrap with 200 resamples, with 2,000 Monte Carlo simulations per resample. Analyses were repeated across 20 multiply imputed datasets, and CIFs and risk differences were combined pointwise across imputations using the mean and empirical 2.5th and 97.5th percentiles. Model adequacy was assessed by comparing natural-course simulated CIFs with nonparametric Aalen–Johansen estimates from the observed data.

#### Technical Details of Multiple Imputation

##### Assumption and overall approach

Missing baseline and time-varying covariates were handled using multiple imputation by chained equations (MICE) under a missing-at-random (MAR) assumption, conditional on variables included in the imputation model. To preserve the longitudinal structure of the data and the causal estimand, outcomes were included as predictors in the imputation models but were not themselves imputed.

##### Pre‑MICE cleaning

Before imputation, physiological and laboratory variables were harmonized and screened for clinical plausibility and structural consistency. Values considered physiologically implausible or ambiguous (for example, inconsistent FiO₂ scales, negative laboratory values, or extreme vital signs) were set to missing rather than corrected or truncated. We did not perform deterministic single imputation or time-series interpolation (for example, last observation carried forward or linear interpolation) before multiple imputation. Structural missingness was handled separately using domain knowledge; for vasopressor administration variables, missing values were interpreted as no drug administration and were recoded to zero, while audit indicators were retained to distinguish structural zeros from observed documentation.

##### Two-phase imputation strategy and variables

We implemented a two-phase imputation pipeline. First, baseline covariates were imputed at the ICU-stay level (Phase 1). Second, daily time-varying covariates were imputed on the person-day dataset (Phase 2), injecting the Phase 1 imputations into each corresponding stay.

The full list of imputed variables, corresponding imputation methods, and their predictive structures are as follows:

- **Phase 1: Baseline Covariates (ICU-stay level)**
  - *Continuous variables (imputed via Predictive Mean Matching [PMM]):* Baseline weight, Height, Albumin, Baseline Hemoglobin.
  - *Log-transformed continuous variables (imputed via PMM and back-transformed):* Baseline Bilirubin, Baseline Creatinine, Baseline Platelets.
  - *Binary variables (imputed via Logistic Regression):* First care unit (MICU indicator).
  - *Predictors used:* Age, Congestive heart failure, Chronic pulmonary disease, Renal disease, Liver disease, Rheumatic disease, plus all Phase 1 imputation targets.
- **Phase 2: Time-Varying Covariates (Person-day level)**
  - *Continuous variables (imputed via PMM):* Daily Hemoglobin, SpO₂, FiO₂, Systolic BP, Diastolic BP.
  - *Log-transformed continuous variables (imputed via PMM and back-transformed):* Bilirubin, Creatinine, Platelets, Daily Urine Output.
  - *Ordinal clinical score (imputed via PMM):* Glasgow Coma Scale (GCS).
  - *Predictor Hierarchy:* To prevent unstable chained predictions among highly missing daily variables, Phase 2 longitudinal targets did not predict one another. Instead, a strict, sparse hierarchical predictor matrix was utilized:
    - **Tier 0 (Core Covariates):** Age, Baseline Hemoglobin.
    - **Time Variable:** Time (Day).
    - **Tier 2 Phase 1 (Baseline Severity Proxies):** Baseline Albumin, Log-transformed Baseline Bilirubin, Log-transformed Baseline Creatinine, and Log-transformed Baseline Platelets.
    - **Tier 2 Phase C (Causal Auxiliary Proxies):** Prior-day infection history (when available), Death status, ICU Discharge status, and Dobutamine use.
    - **Variable-specific Extensions:**
      - *Daily Hemoglobin:* Concurrent RBC transfusion status.
      - *FiO₂:* Observed PaO₂ (when available), Blood gas sampling indicator, and Observed SpO₂ (when available).
      - *Creatinine:* Observed daily urine output (when available).
      - *Systolic BP:* Observed Diastolic BP (when available) and Vasopressor rates.
      - *Diastolic BP:* Vasopressor rates.

*Note: For Phase 2 continuous time-varying targets, predictive mean matching (PMM) with 5 donors was used exclusively rather than parametric two-level models (2l.pan). This choice was made to strictly preserve clinical bounds and prevent negative or out-of-range draws for physiological variables,* t*o preserve clinically plausible values, although this approach does not explicitly model within-stay clustering in the conditional imputation models.*

##### Implementation details, diagnostics, and pooling

Multiple imputation by chained equations was performed with 20 imputations (m = 20) and up to 10 iterations per imputed dataset. Convergence was achieved by iteration 6. Imputation was conducted in R (version 4.3.1) using the mice package (version 3.17.144) with additional multilevel functionality (pan and lme4 backends for two-level methods).

### Supplementary Table 6. Strategy-Specific Transfusion Summary (MI-Pooled)

| Metric | Restrictive | Liberal | No-transfusion |
| --- | --- | --- | --- |
| **Original Patients N** | 4013 | 4013 | 4013 |
| **Active Clones** | 4013 | 4013 | 4013 |
| **Person-days (mean [min-max])** | 19457 [19425 ‒ 19504] | 16161 [16101 ‒ 16203] | 19303 [19303 ‒ 19303] |
| **Censoring due to Deviation % (Pooled ± 95%CI)** | 16.3% (15.2% ‒ 17.5%) | 29.1% (27.7% ‒ 30.5%) | 16.8% (15.6% ‒ 17.9%) |
| **Transfused Patients % (Pooled ± 95%CI)** | 1.1% (0.7% ‒ 1.4%) | 8.0% (7.1% ‒ 8.8%) | 0.0% (0.0% - 0.0%) |
| **Total RBC Volume mL (mean [min-max])** | 49985 [45974 ‒ 53738] | 376474 [375399 ‒ 377924] | 0.0 [0.0 - 0.0] |
| **Mean RBC per Transfused Patient mL (Pooled ± 95%CI)** | 699.1 (564.3 ‒ 833.8) | 865.2 (792.7 ‒ 937.7) | 0.0 (0.0 - 0.0) |

*Note: Descriptive statistics were calculated using person-days during which clones remained adherent to the assigned strategy. Results were pooled across 20 multiply imputed datasets.*

### Supplementary Figure 3. Daily Transfusion Proportion

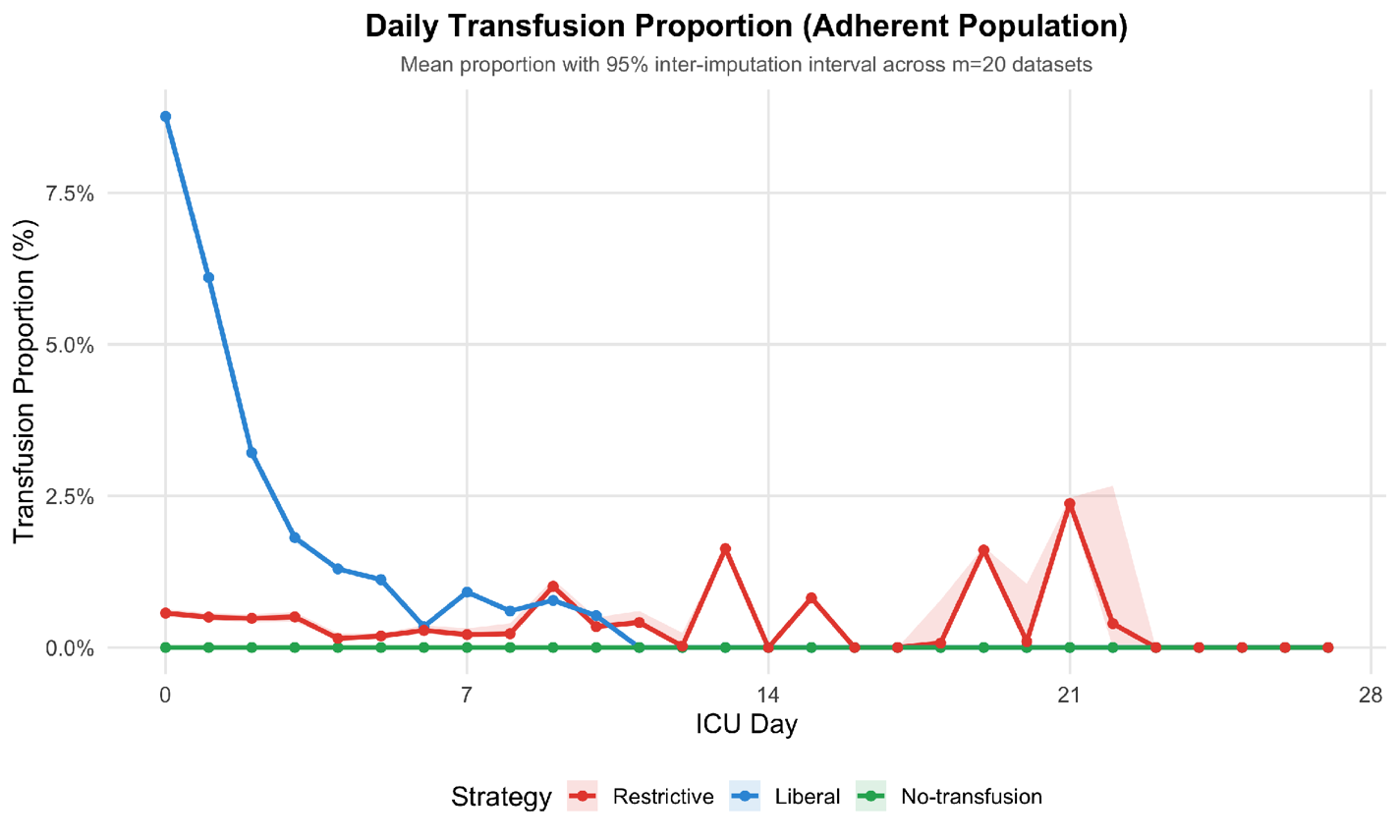

*Daily Transfusion Proportion*

*Figure note: Mean daily transfusion proportion with 95% inter-imputation interval across 20 multiply imputed datasets within the adherent population.*

### Supplementary Figure 4. Cumulative Transfusion Volume (Among Transfused)

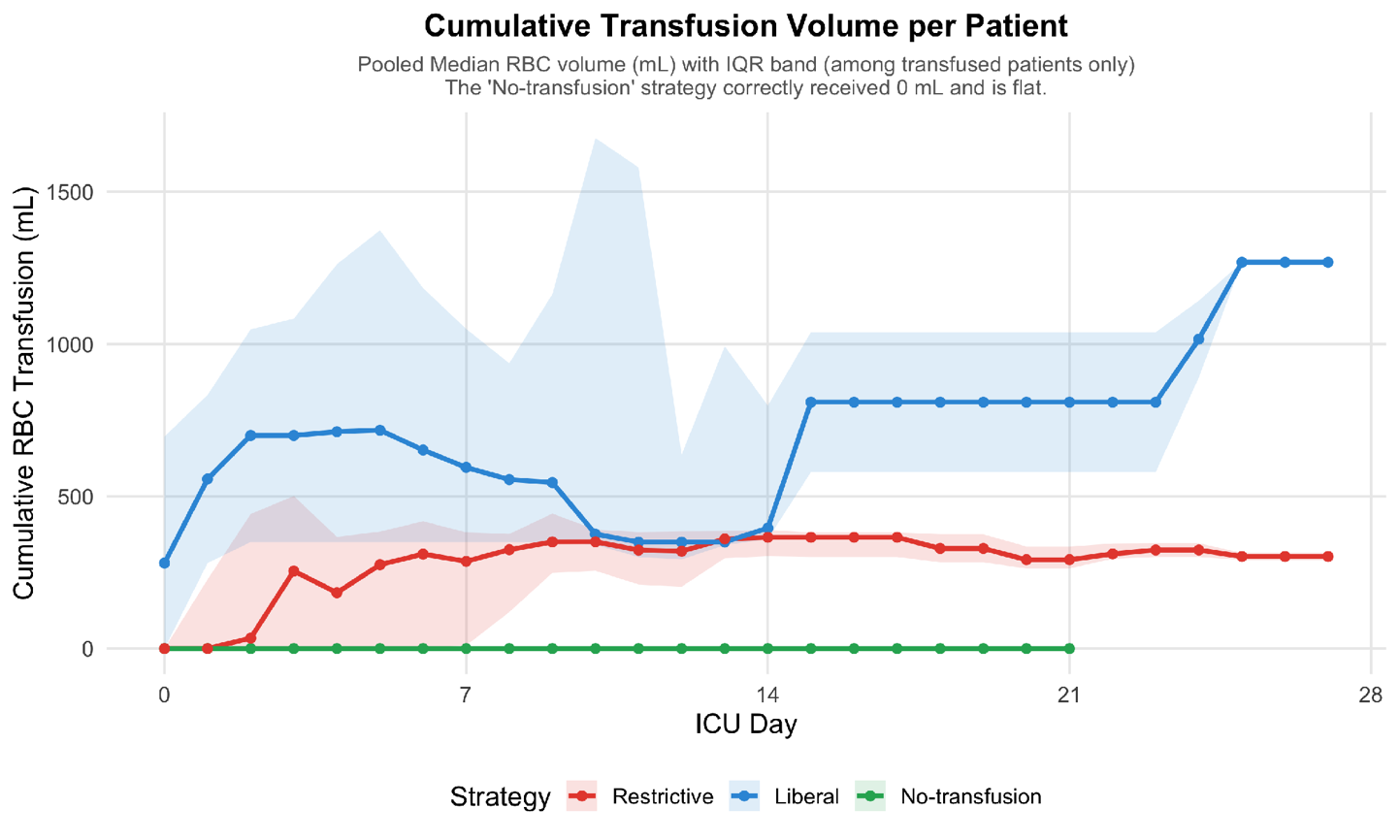

*Cumulative Transfusion Volume*

*Figure note: Pooled median cumulative RBC volume (mL) with interquartile range (IQR) band among transfused patients. The ‘No-transfusion’ strategy correctly received 0 mL.*

### Supplementary Table 7. Subgroup Analysis: SOFA > 5 — Weight Diagnostics, Effective Sample Size, and MSM Estimates

This section presents the results of a pre-specified subgroup analysis restricted to patients with a baseline SOFA score > 5 (N = 2,568 of 4,013; 64.0%). The clone-censor-weight (CCW) pipeline and marginal structural model (MSM) were applied identically to the primary analysis, with baseline covariates (L0) included in both the IPCW numerator and the MSM outcome model. All estimates were pooled across m = 20 multiply imputed datasets using Rubin’s rules.

*Panel A. Weight Diagnostics*

Distribution of stabilized IPCW weights at the primary truncation threshold (97.5th percentile). Values represent means across 20 imputations. ESS = Effective Sample Size, computed as (ΣW)² / ΣW².

| Mean | SD | Median | 95th Percentile | Maximum | ESS Ratio |
| --- | --- | --- | --- | --- | --- |
| 0.997 | 0.092 | 0.998 | 1.167 | 1.265 | 99.2% |

The stabilized IPCW weights were well-behaved: the mean weight remained close to 1.0, the maximum weight was 1.27 after truncation, and the ESS ratio exceeded 99%. These diagnostics indicate no evidence of positivity violations or extreme weight instability in this subgroup.

*Panel B. Effect Estimates*

Adjusted effect estimates for ICU-acquired infection, with the Restrictive strategy as the reference group. Estimates are shown for the primary truncation threshold (97.5th percentile).

| Comparison | Measure | Estimate | 95% CI |
| --- | --- | --- | --- |
| Liberal vs Restrictive | Conditional OR | 0.941 | [0.767, 1.154] |
|  | Marginal OR | 0.941 | [0.770, 1.152] |
|  | Marginal RD (pp) | −0.04 | [−0.18, 0.10] |
| No-transfusion vs Restrictive | Conditional OR | 0.995 | [0.927, 1.069] |

Estimates were similar when stabilized weights were truncated at the 95th percentile (conditional OR, 0.939; 95% CI, 0.766–1.151) and the 99th percentile (conditional OR, 0.942; 95% CI, 0.767–1.156). Marginal effect estimates were equally stable across thresholds (marginal OR range, 0.939–0.942; marginal RD, −0.04 percentage points for all thresholds).

### Supplementary Table 8. Sensitivity Analyses: Causal Effect of Transfusion Strategies on ICU-Acquired Infection

This table summarizes the pooled odds ratios (ORs), risk differences (RDs), and 95% confidence intervals (CIs) from the marginal structural models across multiple pre-specified sensitivity analyses. The Restrictive strategy serves as the reference group for all comparisons. The primary analysis includes baseline covariates (L0) in both the IPCW numerator and the MSM outcome model, following Cole and Hernán’s consistency framework.

| Analysis | Liberal Strategy (Conditional OR) | P-value | No-transfusion Strategy (Conditional OR) | P-value | Liberal Strategy (Marginal OR) | P-value |
| --- | --- | --- | --- | --- | --- | --- |
| Primary | 0.954 [0.797, 1.142] | 0.609 | 0.994 [0.926, 1.067] | 0.864 | 0.955 [0.800, 1.139] | 0.608 |
| No baseline vars | 1.023 [0.787, 1.330] | 0.865 | 0.996 [0.861, 1.153] | 0.960 | 1.023 [0.789, 1.326] | 0.865 |
| Deviation Timing | 1.056 [0.830, 1.342] | 0.658 | 0.951 [0.885, 1.022] | 0.171 | 1.055 [0.835, 1.333] | 0.655 |
| Landmark Day 1 | 0.972 [0.817, 1.157] | 0.750 | 0.984 [0.926, 1.045] | 0.603 | 0.972 [0.818, 1.155] | 0.751 |
| Truncation 95% | 0.943 [0.791, 1.124] | 0.512 | 0.987 [0.932, 1.045] | 0.647 | 0.944 [0.795, 1.120] | 0.507 |
| Truncation 99% | 0.948 [0.793, 1.134] | 0.560 | 0.986 [0.925, 1.052] | 0.675 | 0.949 [0.797, 1.130] | 0.556 |
| Complete Case* | 1.333 [1.333, 1.333] | 0.000 | 1.003 [1.003, 1.003] | 0.000 | 1.000 [1.000, 1.000] | 0.558 |
| 28-Day HAI Observation | 1.018 [0.881, 1.175] | 0.809 | 0.979 [0.929, 1.031] | 0.413 | 1.018 [0.883, 1.173] | 0.807 |

*Note: The complete case analysis excluded all patient-stays with missing values in any required baseline or time-varying covariate during the at-risk period, leaving an effective sample size of 111 stays. Results represent conditional odds ratios from the pooled unweighted model due to sample size constraints for marginal standardization.*

### Supplementary Table 9. 28-Day Cumulative Infection Risk (Pooled, m = 20)

| Strategy | Risk | 95% CI | RD (vs Restrictive) | 95% CI |
| --- | --- | --- | --- | --- |
| Restrictive | 2.16% | 2.07 – 2.25 | Reference | — |
| Liberal | 2.14% | 2.06 – 2.23 | −0.02% | −0.15 – 0.11 |
| No-transfusion | 2.22% | 2.12 – 2.32 | +0.06% | −0.08 – 0.20 |
| Natural course | 2.14% | 2.05 – 2.23 | −0.02% | −0.15 – 0.11 |

Risk differences are expressed as percentage-point differences.

### Supplementary Table 10. Day 28 Outcome Distribution (Mean ± SD across m = 20 imputations)

| Strategy | Infection (%) | Death (%) | Discharge (%) | Event-Free (%) |
| --- | --- | --- | --- | --- |
| Restrictive | 2.16 ± 0.19 | 12.16 ± 0.42 | 85.52 ± 0.52 | 0.16 ± 0.04 |
| Liberal | 2.14 ± 0.18 | 11.67 ± 0.41 | 85.98 ± 0.47 | 0.20 ± 0.06 |
| No-transfusion | 2.22 ± 0.21 | 12.08 ± 0.45 | 85.53 ± 0.45 | 0.17 ± 0.05 |
| Natural course | 2.14 ± 0.19 | 11.98 ± 0.57 | 85.69 ± 0.60 | 0.19 ± 0.05 |

**Definition of the Natural Course**: Under the natural course, no hypothetical transfusion intervention was imposed, and transfusion and subsequent time-varying processes were simulated using models fitted to the observed data. This scenario was distinct from the no-transfusion strategy, under which RBC transfusion was set to zero throughout follow-up. The simulated 28-day cumulative incidence of ICU-acquired infection under the natural course was compared with the corresponding Aalen–Johansen estimate from the observed data. Across all 20 multiply imputed datasets, the absolute differences were within the prespecified tolerance margin, with both mean estimates being approximately 2.14%.

### Supplementary Figure 5. Death Cumulative Incidence Function (CIF) by Transfusion Strategy

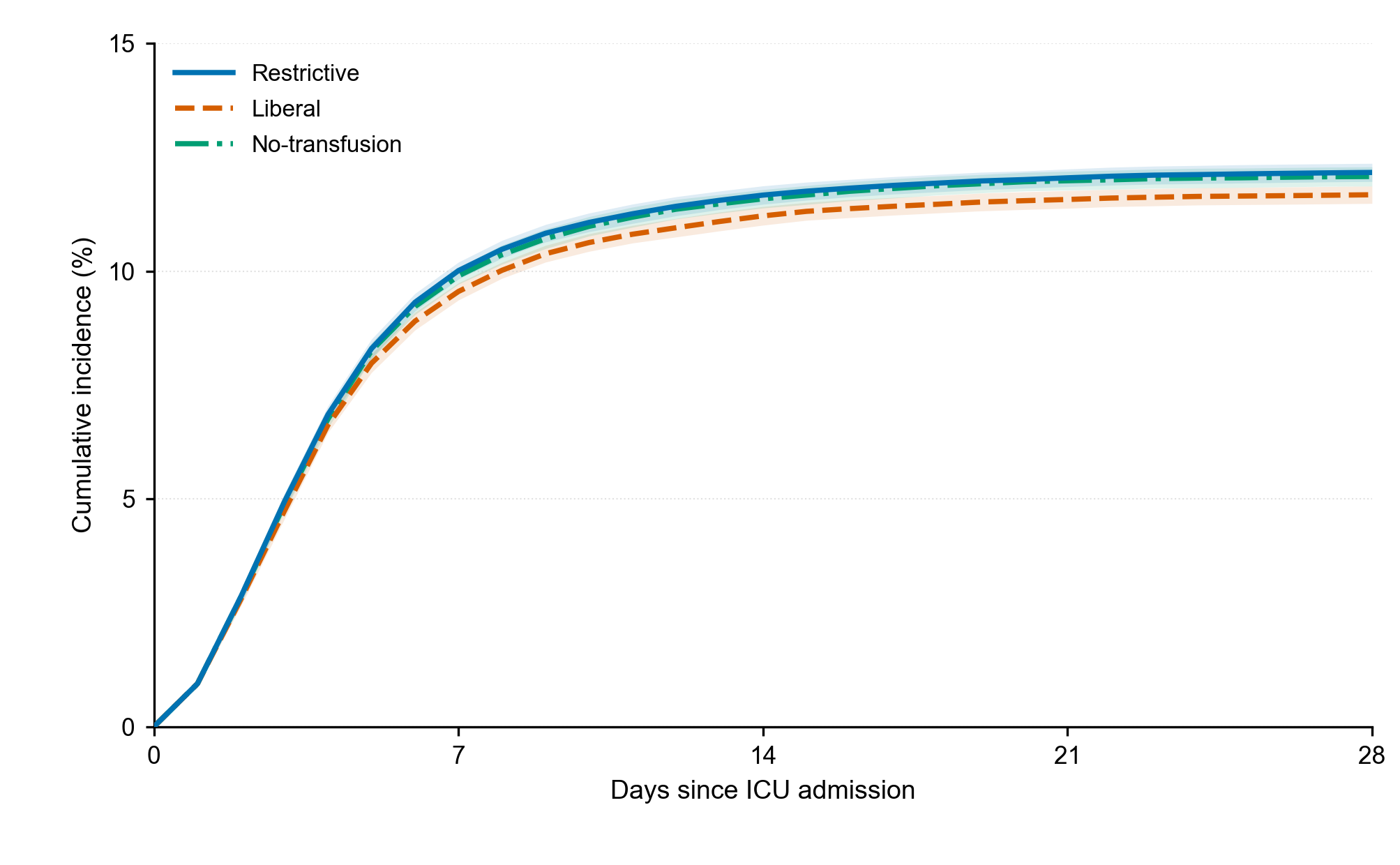

Figure note: Cumulative incidence of in-ICU death by transfusion strategy.

### Supplementary Figure 6. Discharge Cumulative Incidence Function (CIF) by Transfusion Strategy

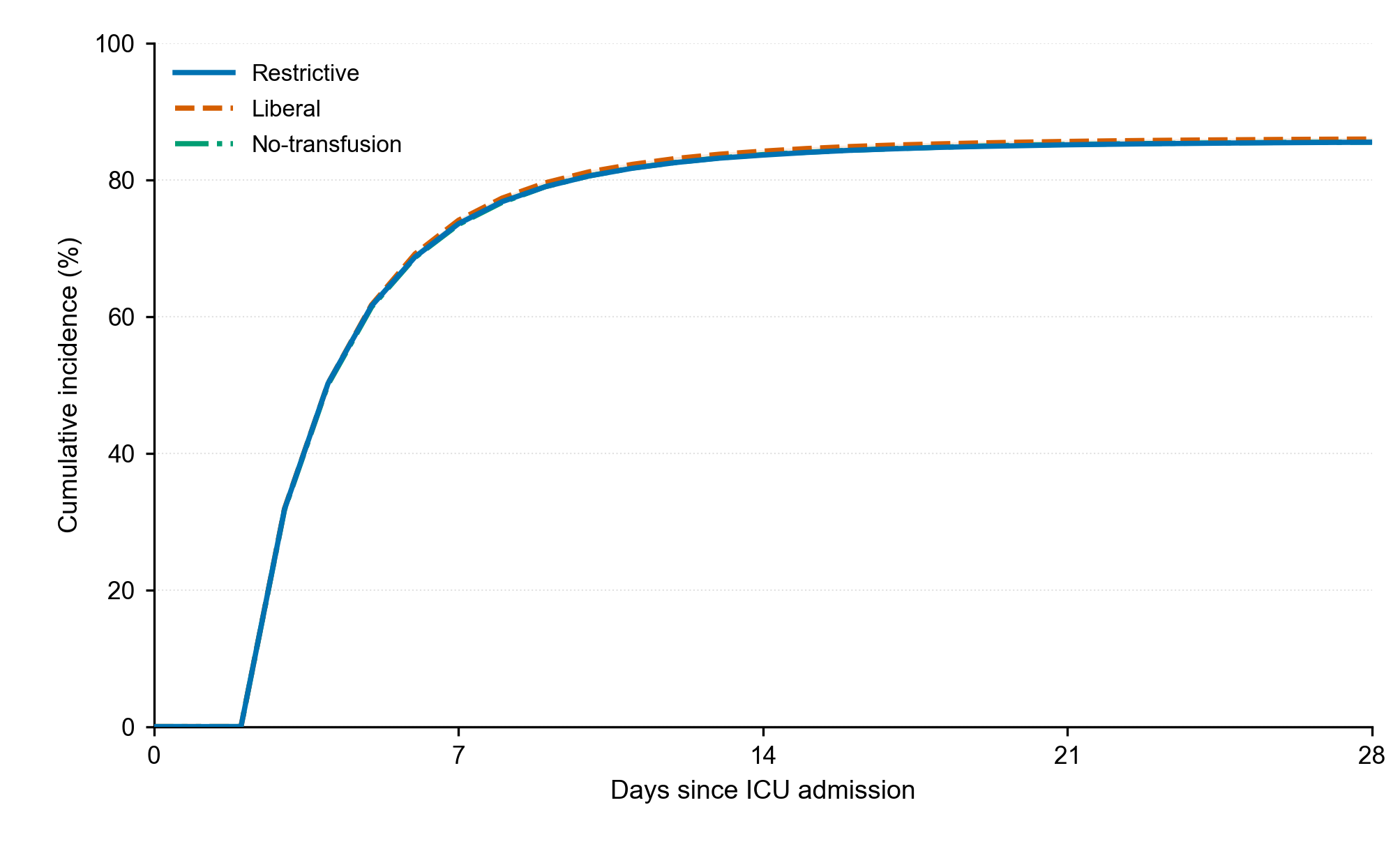

Figure note: Cumulative incidence of ICU discharge by transfusion strategy.
